# Multimodal Molecular Imaging Reveals Tissue-Based T Cell Activation and Viral RNA Persistence for Up to 2 Years Following COVID-19

**DOI:** 10.1101/2023.07.27.23293177

**Authors:** Michael J. Peluso, Dylan Ryder, Robert Flavell, Yingbing Wang, Jelena Levi, Brian H. LaFranchi, Tyler-Marie Deveau, Amanda M. Buck, Sadie E. Munter, Kofi A. Asare, Maya Aslam, Wally Koch, Gyula Szabo, Rebecca Hoh, Monika Deswal, Antonio Rodriguez, Melissa Buitrago, Viva Tai, Uttam Shrestha, Scott Lu, Sarah A. Goldberg, Thomas Dalhuisen, Matthew S. Durstenfeld, Priscilla Y. Hsue, J. Daniel Kelly, Nitasha Kumar, Jeffrey N. Martin, Aruna Gambir, Ma Somsouk, Youngho Seo, Steven G. Deeks, Zoltan G. Laszik, Henry F. VanBrocklin, Timothy J. Henrich

**Affiliations:** Division of HIV, Infectious Diseases, and Global Medicine, University of California San Francisco, San Francisco, CA USA; Division of Experimental Medicine, University of California San Francisco; Department of Radiology and Biomedical Imaging, University of California San Francisco; CellSight Technologies, San Francisco, CA; Department of Pathology, University of California San Francisco; Department of Epidemiology and Biostatistics, University of California San Francisco; Division of Cardiology, University of California San Francisco; Division of Gastroenterology, University of California San Francisco

**Keywords:** COVID-19, SARS-CoV-2, post-acute sequelae of SARS-CoV-2 (PASC), Long COVID, PET imaging, viral persistence, immune activation

## Abstract

The etiologic mechanisms of post-acute medical morbidities and unexplained symptoms (Long COVID) following SARS-CoV-2 infection are incompletely understood. There is growing evidence that viral persistence and immune dysregulation may play a major role. We performed whole-body positron emission tomography (PET) imaging in a cohort of 24 participants at time points ranging from 27 to 910 days following acute SARS-CoV-2 infection using a novel radiopharmaceutical agent, [^18^F]F-AraG, a highly selective tracer that allows for anatomical quantitation of activated T lymphocytes. Tracer uptake in the post-acute COVID group, which included those with and without Long COVID symptoms, was significantly higher compared to pre-pandemic controls in many anatomical regions, including the brain stem, spinal cord, bone marrow, nasopharyngeal and hilar lymphoid tissue, cardiopulmonary tissues, and gut wall. Although T cell activation tended to be higher in participants imaged closer to the time of the acute illness, tracer uptake was increased in participants imaged up to 2.5 years following SARS-CoV-2 infection. We observed that T cell activation in spinal cord and gut wall was associated with the presence of Long COVID symptoms. In addition, tracer uptake in lung tissue was higher in those with persistent pulmonary symptoms. Notably, increased T cell activation in these tissues was also observed in many individuals without Long COVID. Given the high [^18^F]F-AraG uptake detected in the gut, we obtained colorectal tissue for *in situ* hybridization SARS-CoV-2 RNA and immunohistochemical studies in a subset of participants with Long COVID symptoms. We identified cellular SARS-CoV-2 RNA in rectosigmoid lamina propria tissue in all these participants, ranging from 158 to 676 days following initial COVID-19 illness, suggesting that tissue viral persistence could be associated with long-term immunological perturbations.

## INTRODUCTION

Some people do not return to their baseline health following SARS-CoV-2 infection^1, 2^. Following the acute phase, such individuals may experience an increased burden of new onset medical conditions such as cardiovascular disease or diabetes mellitus^3^. They may also experience Long COVID (LC), defined as unexplained symptoms or changes in health not attributable to an alternative diagnosis^2^. The U.S. Centers for Disease Control and Prevention (CDC) recently reported that approximately 15% of American adults have experienced Long COVID at any time and that 6% are currently experiencing the condition^4^; up to 18 million adults in the U.S. alone might be affected^5^. Despite the scale of the problem, there are currently no accepted treatments and massive efforts are now underway to understand the pathophysiology of these post-acute sequelae, including Long COVID^6, 7^.

Acute COVID-19 is a highly inflammatory illness^8, 9^. In the post-acute phase, inflammation, immune activation and long-term dysregulation of virus-specific immune responses have consistently been identified in peripheral blood^10–22^. These immune responses have been associated with a variety of factors including clotting dysfunction,^23–26^ reactivation of latent viral infections such as Epstein Barr Virus (EBV),^10, 27, 28^ and autoimmune responses^10, 29–33^. Importantly, there is a growing body of evidence that persistent SARS-CoV-2 RNA or protein can be detected in various tissue compartments for many months following acute infection^34–41^. This may explain, at least in part, ongoing aberrant immune responses, inflammation, and clinical symptomatology^42, 43^.

Despite advances in understanding systemic inflammation in Long COVID, data regarding the role of SARS-CoV-2 persistence or aberrant T cell responses in non-blood tissues are sparse. Most studies to date have been limited to small autopsy or biopsy samples from convenience cohorts^35, 38^, with many individuals requiring hospitalization during acute infection or without detailed data on the post-acute course. Clinical studies that have evaluated tissue pathology in living participants have assessed limited quantities of tissue obtained through minimally invasive clinical biopsies. Furthermore, anatomic regions such as brain, spinal cord, cardiopulmonary tissue, vascular tissue, and other potential sites of SARS-CoV-2 persistence cannot be sampled in living individuals via biopsy procedures^44, 45^. As a result, characterization of the immune responses in these anatomical locations, including processes like T cell activation and trafficking, has been limited. When it has been attempted, it has utilized non-specific tracers or limited follow-up of clinical symptoms^46–53^. Therefore, there is an urgent need to develop non-invasive techniques to identify more specific persistent and/or aberrant immune responses in highly characterized cohorts over the long-term to better understand the tissue-level biology that might drive findings observed in peripheral blood.

In this study, we performed whole-body positron emission tomography (PET) imaging in 24 highly characterized participants from the UCSF-based LIINC cohort (NCT04362150)^54^ at time points ranging from 27 to 910 days following COVID-19 symptom onset. We used a novel radiopharmaceutical agent, [18F]F-AraG (Fluorine-18 labeled arabino furanosyl guanine), a selective and sensitive tracer that allows for anatomical localization of activated CD8+ and CD4+ T lymphocytes^55–57^. We found that [^18^F]F-AraG uptake was significantly higher in many anatomical regions among post-acute COVID participants compared to pre-pandemic controls. These included the brain stem, spinal cord, bone marrow, nasopharyngeal and hilar lymphoid tissue, cardiopulmonary tissues, and gut wall. Increased uptake was identified up to 2.5 years following SARS-CoV-2 infection in the absence of known re-infection. Furthermore, [^18^F]F-AraG uptake in some tissues was associated with a higher number of Long COVID symptoms. Lastly, we identified cellular SARS-CoV-2 RNA in rectosigmoid lamina propria tissue in all participants with Long COVID symptoms who underwent biopsy, ranging from 158 to 676 days following initial COVID-19 symptom onset, suggesting that tissue viral persistence could be associated with these immunologic findings.

## RESULTS

### Study cohort

Following local institutional review board and radiation safety committee approvals, [^18^F]F-AraG PET/CT imaging was performed on 24 participants identified from the University of California San Francisco (UCSF)-based Long-term Impact of Infection with Novel Coronavirus (LIINC) study (NCT04362150)^54^. We enrolled two groups of individuals: (1) in the early post-acute phase (<90 days from COVID-19 symptom onset with and without complete recovery (n=3 and n=6, respectively), and (2) in the later post-acute phase (>90 days from COVID-19 symptom onset) with and without complete recovery (n=3 who recovered quickly and n=15 with Long COVID symptoms (**Table 1**). In addition, images obtained from 6 participants who underwent [^18^F]F-AraG PET imaging (3 females and 3 males) prior to 2020 were used as pre-pandemic controls. We elected to use pre-pandemic rather than contemporaneous comparators to avoid misclassification of such individuals, given the high rate of subclinical or undiagnosed SARS-CoV-2 infection during the study period.

**Table 1.** Participant Demographics, Clinical Factors and Long COVID History.

| Pt # | Variant Wave at Time of Infection | Hosp. During Acute Infection | Days from Infection to PET Scan<sup>a</sup> | Days From Vaccine to PET | Sex | Age<sup>b</sup> | LC Symptom Count<sup>c</sup> | Presence of LC Symptom Phenotype at Time of Imaging |  |  |  |  |  | Anti-N Ab at PET<sup>d</sup> | Days from COVID to Gut Biopsy<sup>e</sup> |
| --- | --- | --- | --- | --- | --- | --- | --- | --- | --- | --- | --- | --- | --- | --- | --- |
|  |  |  |  |  |  |  |  | Fatigue | Pulm | Cardiac | URI | GI | Neuro |  |  |
| 1 | Pre-Omicron | N | 27 | 135 | F | 25-29 | 2 | Y | Y | N | N | N | N | Y | - |
| 2 | Pre-Omicron | N | 29 | 113 | M | 35-39 | 1 | N | Y | N | N | N | N | Y | - |
| 3 | Pre-Omicron | N | 42 | 151 | M | 30-34 | 4 | N | Y | Y | N | N | N | Y | - |
| 4 | Pre-Omicron | N | 44 | 149 | M | 60-64 | 0 | N | N | N | N | N | N | Y | - |
| 5 | Pre-Omicron | N | 48 | 199 | M | 25-29 | 5 | Y | N | Y | N | Y | Y | Y | - |
| 6 | Pre-Omicron | N | 50 | N/A<sup>f</sup> | F | 30-34 | 4 | Y | Y | N | N | N | Y | N | - |
| 7 | Omicron | N | 64 | 6 | M | 40-44 | 14 | Y | Y | N | N | Y | Y | N | - |
| 8 | Omicron | N | 65 | 386 | F | 60-64 | 0 | N | N | N | N | N | N | N | - |
| 9 | Omicron | N | 83 | 183 | F | 55-59 | 0 | N | N | N | N | N | N | Y | - |
| 10 | Omicron | N | 95 | 246 | M | 45-49 | 0 | N | N | N | N | N | N | Y | 158 |
| 11 | Omicron | N | 114 | 286 | F | 30-34 | 8 | Y | N | Y | Y | Y | N | Y | - |
| 12 | Pre-Omicron | N | 193 | 137 | F | 25-29 | 13 | Y | Y | N | N | Y | Y | N | - |
| 13 | Omicron | N | 205 | 122 | F | 45-49 | 6 | Y | Y | N | N | N | Y | N | - |
| 14 | Omicron | N | 231 | 63 | M | 35-39 | 4 | Y | N | Y | N | Y | Y | Y | - |
| 15 | Omicron | N | 246 | 302 | M | 65-69 | 6 | Y | N | N | Y | Y | Y | N | 213 |
| 16 | Pre-Omicron | Y<sup>g</sup> | 260 | 118 | M | 50-54 | 8 | Y | Y | N | Y | N | Y | N | 442 |
| 17 | Omicron<sup>h</sup> | N | 406 | 219 | F | 30-34 | 0 | N | N | N | N | N | N | Y | - |
| 18 | Pre-Omicron | N | 617 | 320 | F | 30-34 | 7 | Y | N | Y | N | N | Y | N | 645 |
| 19 | Pre-Omicron | N | 625 | 122 | M | 45-49 | 14 | Y | Y | N | N | Y | Y | Y | - |
| 20 | Pre-Omicron | N | 641 | 321 | M | 25-29 | 11 | Y | N | Y | N | Y | Y | Y | - |
| 21 | Pre-Omicron | N | 654 | 425 | F | 30-34 | 15 | Y | Y | Y | N | Y | Y | N | - |
| 22 | Pre-Omicron | N | 663 | 248 | M | 50-54 | 6 | Y | Y | N | Y | Y | Y | Y | 676 |
| 23 | Pre-Omicron | N | 890 | 211 | M | 50-54 | 0 | N | N | N | N | N | N | Y | - |
| 24 | Pre-Omicron | Y<sup>i</sup> | 910 | 155 | F | 55-59 | 6 | Y | Y | N | N | Y | Y | N | - |

Pt = participant; Hosp. = hospitalized; Pulm = pulmonary; URI = upper respiratory; GI = gastrointestinal; Neuro = neurocognitive; LC = Long COVID; Y = Yes, N = No; F = female; M = male
<sup>a</sup> Days from last COVID-19 vaccine dose to PET imaging
<sup>b</sup> Age in years in 5-year age bracket
<sup>c</sup> Number of participant reported symptoms at the time of PET imaging (out of 32 total)
<sup>d</sup> Anti-Nucleocapsid antibody detected at time of PET
<sup>e</sup> All participants had a least one Long COVID symptoms at the time of sigmoidoscopy and rectosigmoid tissue collection
<sup>f</sup> Participant was not vaccinated prior to PET imaging
<sup>g</sup> Participant did not require intensive care but received supplemental oxygen during hospitalization
<sup>h</sup> Participant initial infected during pre-Omicron wave but had two documented re-infections. Days from infection to PET scan calculated from last Omicron reinfection

**Table 1** lists participant demographics, clinical factors and Long COVID symptoms present at the time of imaging. Eleven participants were female, a majority were infected prior to emergence of Omicron variants, and only two were hospitalized during the acute phase of SARS-CoV-2 infection; one had required supplemental oxygen, but not intensive care or mechanical ventilation and the other had not required supplemental oxygen or intensive care. The median age was 39.5 years (range 26 to 65), median number of Long COVID symptoms at the PET screening visit was 5.5 (range 0 to 15), and median number of days from initial COVID-19 symptom onset and PET imaging was 199 (range 27 to 910). The most common symptoms were fatigue (n=16) and neurocognitive complaints (n=14). Six participants did not report any Long COVID symptoms at the time of imaging. All but one participant had received at least one COVID-19 vaccination prior to PET imaging (median number of days from the most recent vaccine dose to tracer injection was 183 days).

To minimize the impact of vaccination on T cell activation, PET imaging was performed greater than 60 days from any vaccine dose (SARS-CoV-2 or otherwise), but one participant received a SARS-CoV-2 booster vaccine dose 6 days prior to imaging without notifying the study team. One participant (number 17) was initially infected during the ancestral wave but experienced two documented re-infections with presumed Omicron variants prior to PET imaging. Except for participant 17, no one reported acute symptoms suggestive of infection with another virus or reinfection with SARS-CoV-2 reinfection between the initial COVID-19 episode and PET imaging; during the study period none had subsequent positive COVID-19 PCR or antigen results beyond their initial confirmatory test.

### Chest CT findings

Review of the chest CTs demonstrated four participants with mild apical scarring and/or reticulation suggesting mild pulmonary fibrosis, one of which demonstrated a bulla in the right lower lobe. The remainder of the scans were normal except for incidental findings likely not attributable to prior COVID-19 infection (*e.g.,* calcified granulomas) as detailed in **Supplemental Table 1**. These findings are consistent with prior studies demonstrating relatively a relatively low rate of clinically significant fibrotic lung disease detectable by CT in patients with recent SARS-CoV-2 infection^58^.

### [^18^F]F-AraG PET/CT imaging

[^18^F]F-AraG is a novel PET imaging agent developed for the purpose of assessing T cell activation and cycling. It is an analog of arabinosyl guanine (AraG), an FDA-approved chemotherapy drug (nelarabine) used for treatment of refractory malignancies of the T cell lineage^59^. [^18^F]F-AraG can be phosphorylated by cytoplasmic deoxycytidine kinase (dCK) and deoxyguanosine kinase (dGK), two enzymes upregulated in activated T cells, which traps [^18^F]F-AraG intracellularly^60^. The selective uptake of [^18^F]F-AraG by activated T cells has been confirmed *in vivo* using murine models^55, 61^. [^18^F]F-AraG has been shown to be safe with no major adverse events in the context of healthy volunteers, metastatic cancer, and other immune deficiencies^55–57^.

[^18^F]F-AraG was prepared in a two-step method using a modified, previously reported procedure^59^. [^18^F]F-AraG was administered intravenously (166.5-185 MBq) followed by PET/CT (Siemens Biograph Vision) for convalescent COVID-19 participants and without Long COVID symptoms and (299.7-329.3 MBq) followed by PET/MRI (GE SIGNA) for pre-pandemic controls. Whole-body imaging was carried out over an interval approximately 50 minutes post injection, covering regions from vertex to mid-thigh. Regions of interest were drawn around various tissues using isometric 3-dimensional sphere depending on the anatomical structure and the maximum and mean standardized uptake values (SUVmax and SUVmean) were determined. SUVs are a function of the concentration of radioactivity within a ROI, the administered activity, participant weight as a surrogate for volume, and uptake time^62^, allowing cross-participant comparisons of PET activity. Overall, the tracer was well-tolerated, with no serious adverse events reported during and/or following tracer injection and PET/CT imaging. CT was chosen for anatomical localization and PET attenuation correction to provide information on lung parenchymal and structural pathology following COVID infection.

### Increased [^18^F]F-AraG tissue uptake in post-acute COVID participants compared to pre-pandemic controls

As shown in **Figs 1 & 2**, significantly higher SUVmax and SUVmean values were observed across a variety of anatomic regions and tissue types in post-acute COVID participants compared with uninfected controls using two-tailed Kruskal-Wallis tests adjusting for false discovery rates within specific tissue regions (e.g., lymphoid tissues, glandular tissue, vascular, spinal cord, etc.). Maximum Intensity Projections (MIP) of PET data from all post-acute COVID and pre-pandemic control participants are shown in **Supplemental Fig 1**. Although [18F]F-AraG uptake was low overall in brain and spinal cord tissues (*i.e.,* SUVmax and SUVmean <1), significantly higher SUVmax and SUVmean were identified in the thoracic spinal cord and cauda equina (at the level of the fourth lumbar vertebra) and higher SUVmean was identified in the brain stem (pons) as in **Fig 2**. The CNS choroid plexus is known to express high levels of ACE-2, but this region had high background uptake and there were no differences between post-acute COVID cases and pre-pandemic controls. Significantly higher levels of [^18^F]F-AraG uptake (SUVmax and SUVmean) were also observed in the aortic arch, pulmonary artery and lower lung lobes compared with pre-pandemic controls. Significant increases in [^18^F]F-AraG uptake were observed in nasal turbinates (SUVmax and SUVmean), hilar lymph node regions (right-sided; SUVmax), proximal colon wall (SUVmax), rectal wall (SUVmax and SUVmean), lumbar (SUVmax) and iliac crest (SUVmax and SUVmean) bone marrow and pharyngeal tonsils (SUVmax). Although not achieving statistical significance, increased tracer uptake was observed in parotid glands and right heart ventral wall in post-acute COVID participants compared to pre-pandemic controls (**Fig 2**). Uptake in the liver (a metabolic and excretory organ for [18F]F-AraG), abdominal adipose tissue, and quadriceps muscles were similar across all participants. No significant differences in SUV were observed in testes, penile tissue, prostate, or uterine tissue, although sample size was limited for these comparisons (**Supplemental Fig 2**).

**Figure 1.**
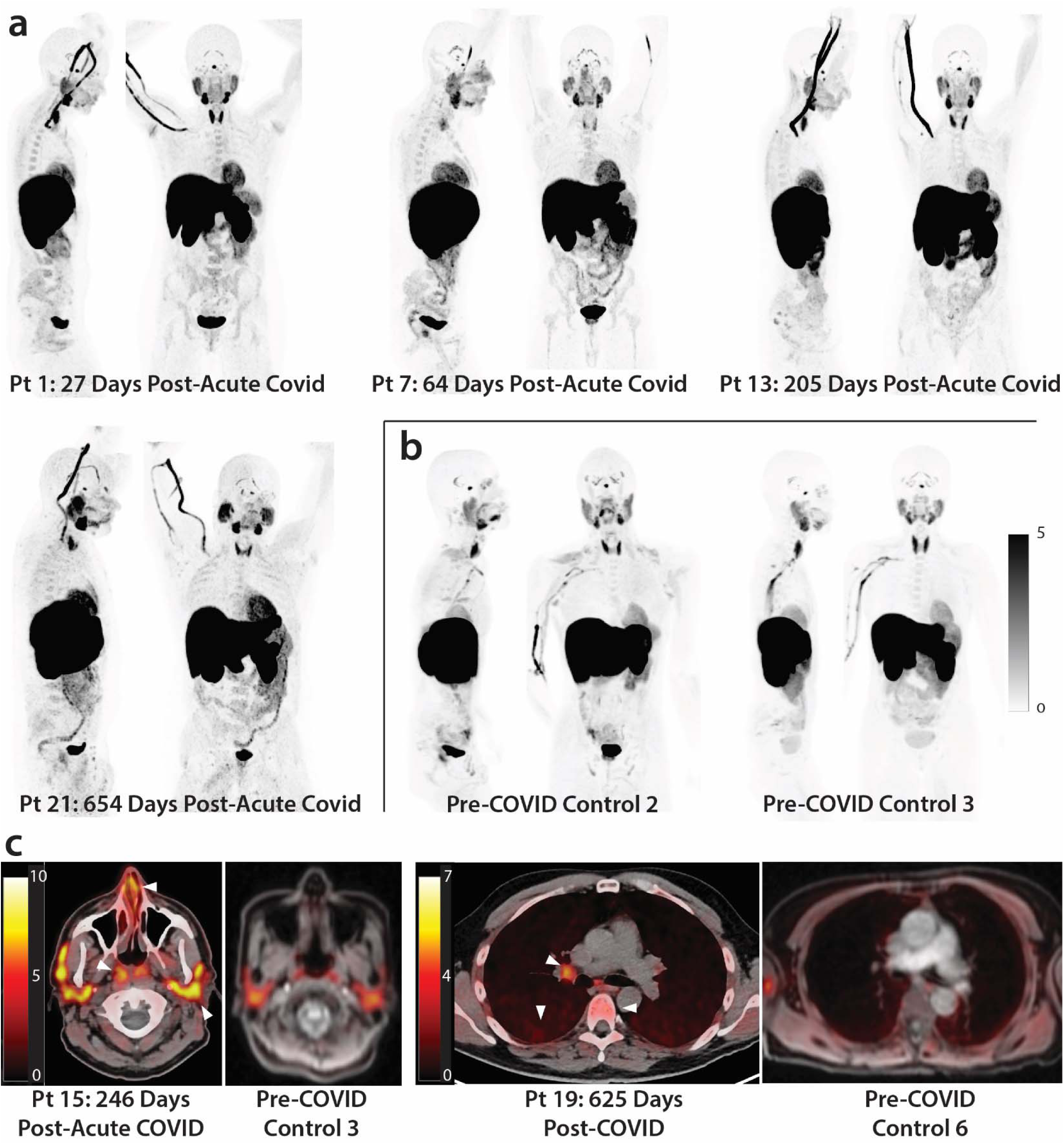
[^18^F]F-AraG PET and PET/CT images from participants following COVID-19 and pre-pandemic control volunteers. Maximum intensity projections (MIP; coronal and sagittal views of 3-dimesional reconstructions) are shown for four representative participants at various times following SARS-CoV-2 infection (**a**) and male and female uninfected controls (**b**). Axial PET/CT overlay images are shown in (**c**) showing increased signal in nasal turbinates, parotid glands, hilar lymph node, lung parenchyma, and lumbar bone marrow in representative post-acute COVID and pre-pandemic control participants (white arrows). MIPs for all participants are shown in **Supplemental Figure 1.**

**Figure 2.**
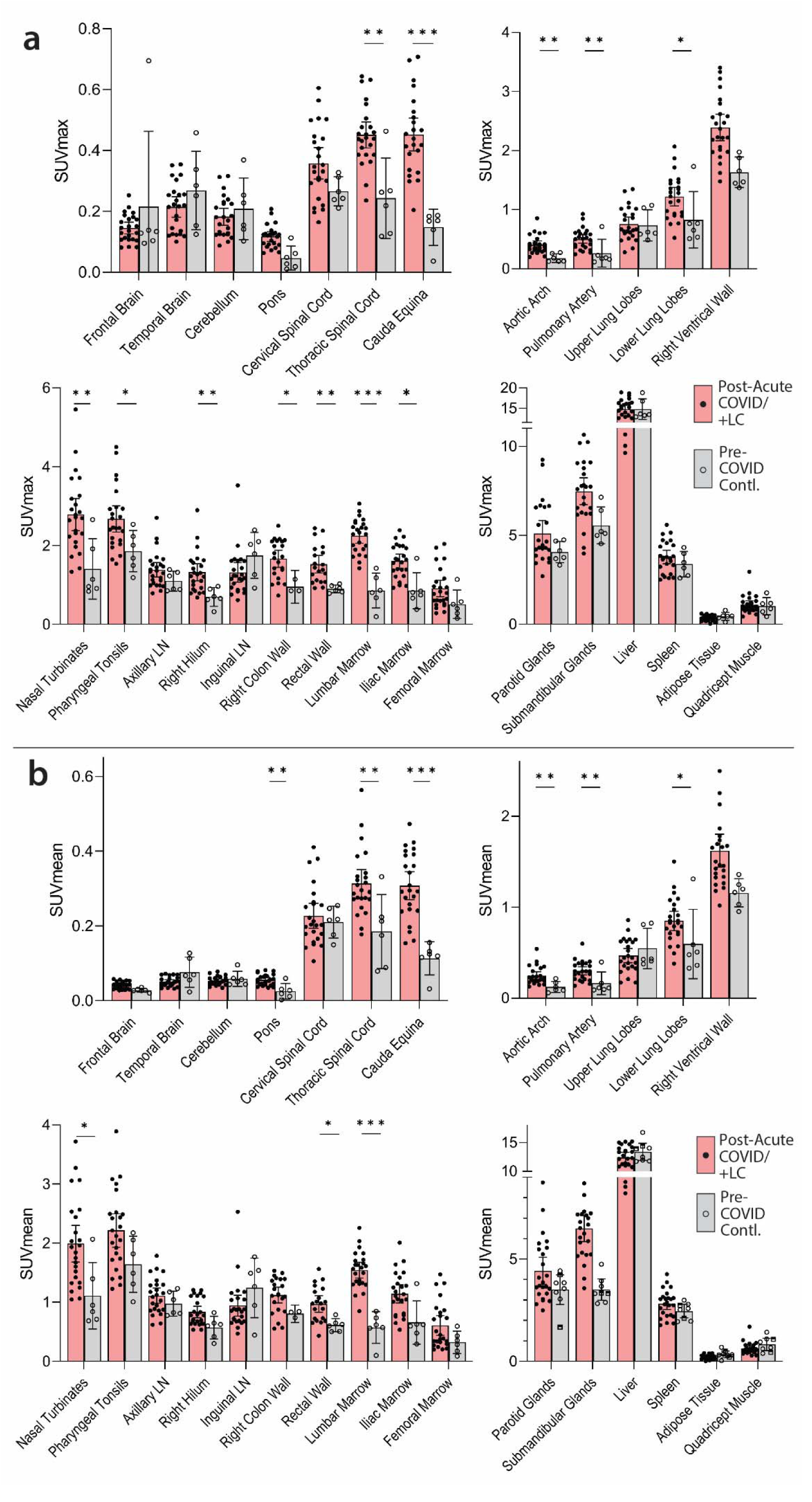
Comparisons of [^18^F]F-AraG standardized uptake values (SUV) in post-acute COVID cases and pre-pandemic control participants. Maximum SUV (SUVmax) (**a**) and mean SUV (SUVmean) (**b**) values for various anatomical regions of interest (ROI) are shown including post-acute COVID participants including those with any number of Long COVID symptoms (Post-acute COVID/+LC; N=18) and pre-pandemic controls (N=6). Bars represent mean SUVmax or SUVmean and error bars represent 95% confidence interval. Adjusted P values <0.05, <0.01 and <0.001 represented by *, **, and *** respectively from two-sided non-parametric Kruskal– Wallis tests using a Benjamini-Hochberg adjustment for false discovery rates across multiple comparisons (q value = adjusted P). All data points are shown on the graph. Gating was not possible in 3 and 5 post-acute COVID participants for proximal colon and rectal wall ROIs, respectively, and 3 pre-pandemic control participants for proximal colon.

### Sex differences in [^18^F]F-AraG tissue uptake

[^18^F]F-AraG uptake grouped by male and female sex is shown in **Supplemental Fig 3.** Although statistical power was limited in these four-way comparison between male and female case and control participants, male participants had trends toward higher tracer uptake in some tissues (pharyngeal tonsils, rectal wall) and significantly higher uptake in hilar ROIs compared to female participants (**Supplemental Fig 3**).

### Impact of time between acute COVID-19 and PET imaging on biodistribution of [^18^F]F-AraG

We performed imaging over a span of nearly two and a half years following COVID-19 symptom onset to determine the duration of T cell activation states in tissues. **Fig 3a** shows [^18^F]F-AraG SUVmax for tissues of interest stratified by timing of PET imaging before or after 90 days following initial COVID-19 symptom onset. We observed modestly decreased uptake in spinal cord and colon/rectal wall ROIs in participants imaged beyond 90 days following COVID-19 symptom onset, but SUVs in these later-imaged individuals remained significantly elevated compared to pre-pandemic controls, with the exception of the right colon wall. When stratified by time since initial COVID-19 symptom onset, [^18^F]F-AraG uptake in the right ventricle wall was significantly higher in post-acute COVID participants compared with pre-pandemic controls (**Fig 3a**). Of note, no significant correlations between [^18^F]F-AraG uptake in any tissue ROI and time from initial infection to PET imaging in those imaged beyond 90 days were observed (all P>0.05 by two-tailed Spearman tests).

**Figure 3.**
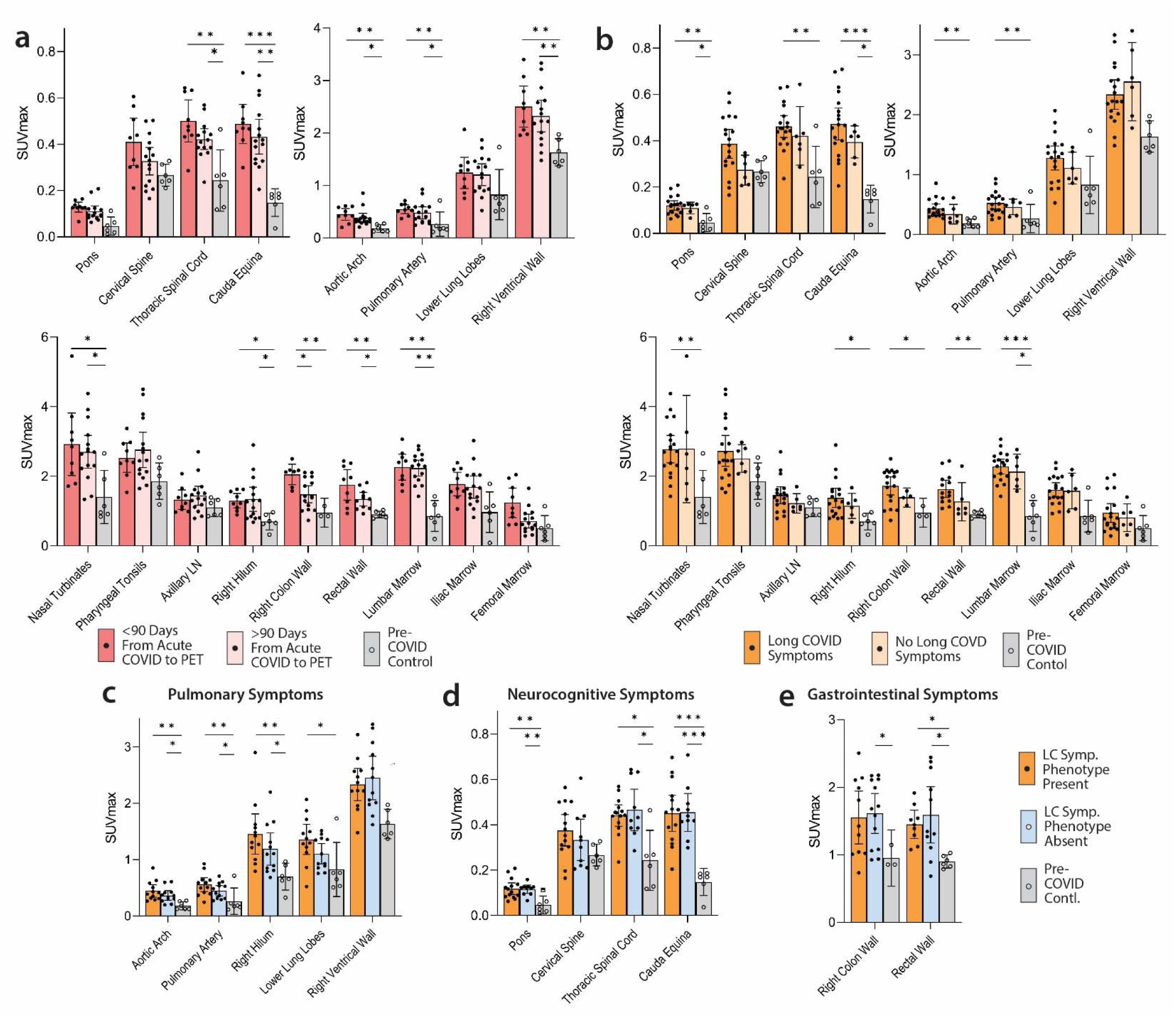
Comparisons of [^18^F]F-AraG maximum standardized uptake values in post-acute COVID cases and control participants grouped by time from initial COVID-19 symptom onset to PET imaging and by Long COVID symptoms. SUVmax values in tissue ROIs in post-acute COVID participants imaged <90 days or =90 days from acute infection onset and control volunteers are shown in (**a**). SUVmax values in tissue ROIs in post-acute COVID participants with or without Long COVID symptoms reported at the time of imaging and control volunteers are shown in (**b**). SUVmax values in tissue ROIs in post-acute COVID participants with or without pulmonary symptoms (c), neurocognitive symptoms (d) and gastrointestinal symptoms (**e**) are also shown. Bars represent mean SUVmax and error bars represent 95% confidence interval. Adjusted P values <0.05, <0.01 and <0.001 represented by *, **, and *** respectively from two-sided non-parametric Kruskal–Wallis tests using a Benjamini-Hochberg adjustment for false discovery rates across multiple comparisons (q value = adjusted P). All data points are shown.

### Long COVID symptoms are associated with higher [^18^F]F-AraG uptake in some tissues

To determine the association between T cell activation and Long COVID-19 symptoms, we compared post-acute COVID participants with (N=18) and without (N=6) Long COVID symptoms at the time of PET imaging. Participants with Long COVID symptoms were generally highly symptomatic with a median 5.5 symptoms reported at the time of imaging. We observed modestly higher uptake in spinal cord, hilar lymph nodes and colon/rectal wall in participants with Long COVID symptoms (**Fig 3b**). To assess relationships between Long COVID symptoms and T cell activity, SUVs in various tissue ROIs were first compared between participants with >5 Long COVID symptoms (N=12) reported at the time of PET imaging, <5 Long COVID symptoms (N=6) and pre-pandemic controls (N=6). SUVmax levels for each group are shown in **Supplementary Fig 4**.

### Association between Long COVID symptom phenotypes and [^18^F]F-AraG tissue uptake

Next, we investigated whether specific Long COVID symptom phenotypes in our cohort correlated with mean SUVmax uptake in tissue ROIs. Participants with pulmonary Long COVID symptoms (e.g., cough, shortness of breath, dyspnea) present at time of imaging had higher SUVmax values in lower lung and hilar ROIs compared to those without pulmonary symptoms (**Figure 3c**). However, direct relationships between other symptom phenotypes (neurocognitive, gastrointestinal, constitutional/fatigue) and tissue uptake in related tissues were not observed. For example, we found similarly increased signal in those with and without specific symptoms phenotypes and pre-pandemic volunteers (representative data shown in **Figure 3d,e**).

### Impact of SARS-CoV-2 vaccination on biodistribution of [^18^F]F-AraG

Since SARS-CoV-2 or other vaccination may impact T cell activation, we analyzed [^18^F]F-AraG uptake in participants grouped by receipt of a SARS-CoV-2 vaccine dose greater than or less than 180 days prior to PET imaging. Timing from most recent vaccination to imaging appeared to have little effect on [^18^F]F-AraG uptake across most tissues, with the exception of modestly lower gut wall tracer uptake in those whose last dose of SARS-CoV-2 vaccine was >180 days prior to imaging (**Supplemental Figure 5a**). Of note, although our protocol excluded those who had received any vaccine within 4 weeks of imaging, one participant received a SARS-CoV-2 mRNA booster 6 days prior to imaging without informing the study team. Post-hoc investigation of this participant’s PET/CT images revealed similar [^18^F]F-AraG uptake across all tissue ROIs to other post-acute COVID participants (*i.e.,* was in the middle range of observed SUVmax) and without marked uptake in the deltoid muscle injection site.

### SARS-CoV-2 antibody responses

SARS-CoV-2 nucleocapsid IgG levels were measured from plasma obtained at the time of PET imaging to provide information about potential recent reinfection that may have influenced our findings. Ten of 24 post-acute COVID participants had no detectable nucleocapsid antibody detection at the time of PET imaging (Signal to Cutoff [SC] ratio <1; **Table 1**). Overall, among the post-acute COVID group, the presence of a detectable nucleocapsid IgG response did not have a major influence on [^18^F]F-AraG uptake across ROIs compared to post-acute COVID (SUVmax values are shown in **Supplemental Figure 5b**). Both those with and without detectable nucleocapsid IgG responses had significantly higher tracer uptake than control volunteers in major ROIs of interest.

### Circulating markers of inflammation and immune activation correlate with [^18^F]F-AraG PET uptake in some tissues

To identify associations between [^18^F]F-AraG PET imaging and levels of systemic inflammation and immune activation, we assessed circulating protein biomarkers in the plasma of participants just prior to the time of PET imaging using the Olink Explore 384 Inflammation panel. Proteomic data were available for 19 of 24 participants. Several differentially expressed gene products were observed in participants grouped by time since initial COVID-19 symptom onset, Long COVID symptom count (proteomic data were only available on two cases without any Long COVID symptoms), and higher PET tracer uptake in various ROIs of interest. Although these did not achieve individual significance after conservative adjustments for multiple comparisons in this small cohort, some interesting patterns were observed. Clustered heatmaps including modules of proteins defined by non-hierarchical k-means clustering by time from initial COVID-19 symptom onset to PET imaging > or ≤90 days, the presence of >5 or ≤5 Long COVID symptoms at the time of imaging, and high or low tracer [^18^F]F-AraG uptake across various anatomical ROIs are shown in **Figure 4**. High and low [^18^F]F-AraG uptake was defined as participants with ROI SUVmax values one to three standard deviations above the mean SUVmax value of the pre-pandemic-19 control subjects (standard deviations cutoffs were based on overall variation within case and control SUVmax values to define clusters of individuals with [^18^F]F-AraG uptake higher than the control population). Time from initial COVID-19 symptom onset to imaging appeared to have a modest impact on differential levels of circulating inflammation markers, with different clusters of differentially expressed gene products in those imaged either <60 or >60 days. In participants reporting >5 Long COVID symptoms at the time of imaging, we observed higher levels of inflammatory markers, including proteins involved in immune responses, chemokine signaling, inflammation responses, and nervous system development (*e.g.* increased expression of proteins such as TGFb1, TANKIL7, TANK, IL20RA, CCL13, SPRY2, PRKAB1, BCR, TAF2). We also observed clusters of gene products upregulated in participants with high levels of lower lung parenchymal [^18^F]F-AraG uptake including those involved in inflammatory response, cell signaling fibroblast transformation, response to mitogenic stimulation among others (*e.g.* differential increased expression of IL-7, CXCL3, CD40, EGF, TRNSF14, TIMP3, CRKl, CXCL3, BKAP2, PDGFB). High parotid gland [^18^F]F-AraG uptake was related to differential expression of gene products involved with matrix metalloprotease, response to bacterial infection, vascular development and coagulation and hypoxic stress as in **Figure 4**. Differential protein expression was more subtle in post-acute COVID participants with high uptake in other tissue ROIs (bone marrow, pharyngeal tonsils, gut wall, nasal turbinates) as in **Supplemental Fig 6**, although there was some evidence for increased differential expression of gene products in post-acute COVID participants with high uptake in the spinal cord which, interestingly, included gene products associated with Alzheimer disease (AGER)^63^, myogenesis/myelination (Cdon^64^), and oxidative stress and monocyte adhesion (MEGF10) protein also involved in mediation of apoptotic cell phagocytosis and amyloid-beta peptide brain uptake and various myopathies^65–67^.

**Figure 4.**
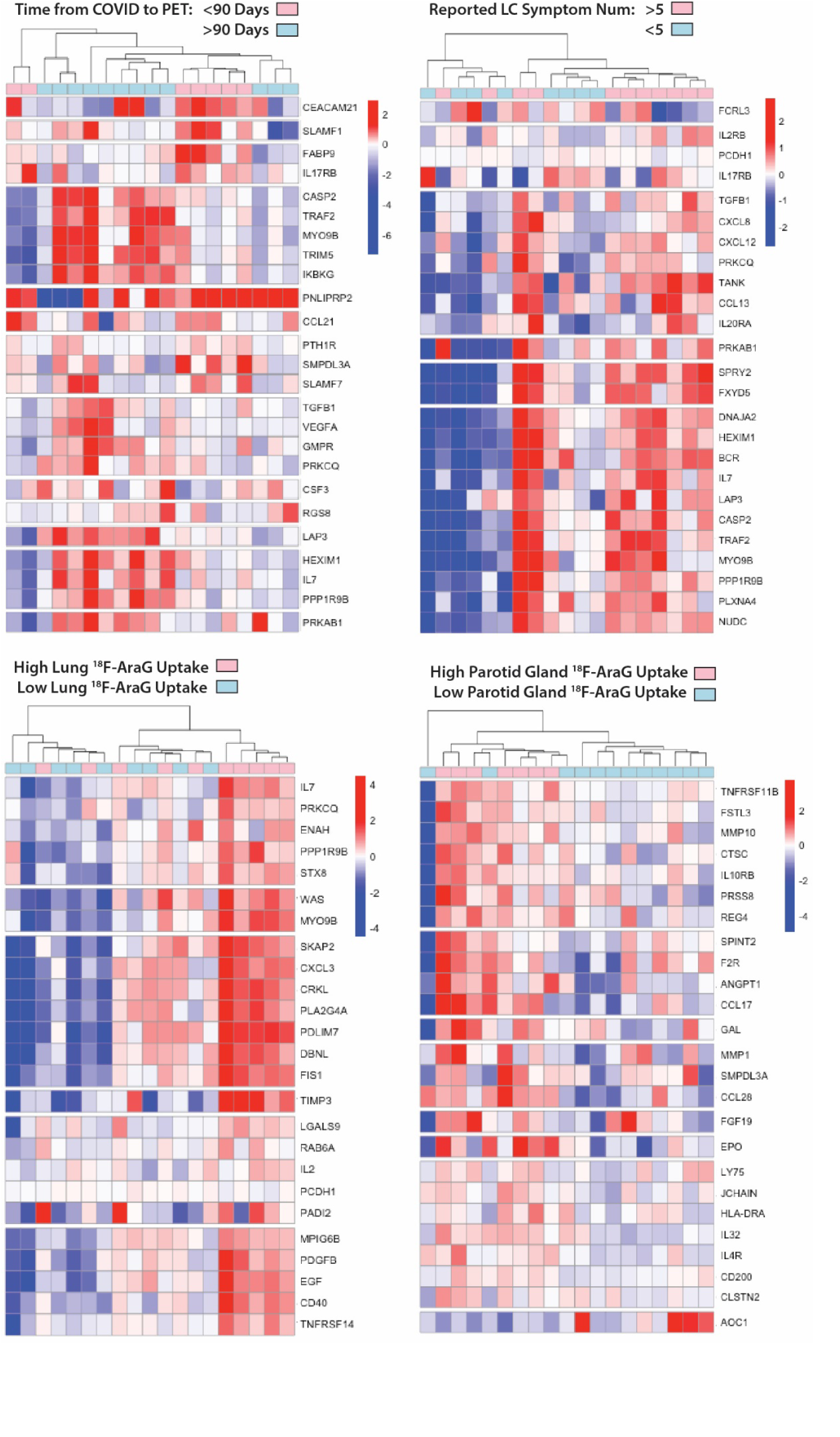
Differential plasma protein expression in post-acute COVID participants grouped by time of COVID to PET imaging, reported Long COVID symptom number and [^18^F]F-AraG uptake in representative tissues. Clustered heat maps of the top 25 differentially expressed plasma proteins from Olink Proximity Extension Assay EXPLORE 384 panel with markers grouped into k-clusters based on similarity are shown for participants imaged early (<90 days) or later (>90 days) after symptom onset (a), those reporting >5 or ≤5 Long COVID symptoms (out of a total of 32 surveyed across multiple organ systems) at the time of PET imaging (b), and in those with high lower lung lobe [^18^F]F-AraG uptake (**c**; defined as SUVmax >2 standard deviations [SD] above the average SUVmax value measured in Pre-pandemic control volunteers), and parotid gland tissue [^18^F]F-AraG uptake (**d**; defined as SUVmax >1 SD above the average SUVmax value measured in pre-pandemic control participants). Note that modules of various gene products representing inflammation and immune activation have higher differential expression in participants with >5 Long COVID symptoms and with higher [^18^F]F-AraG uptake in tissue. Heat maps clustered by high uptake in other tissues ROIs were less revealing and shown in **Supplemental Figure 4**)

**Figure 5.**
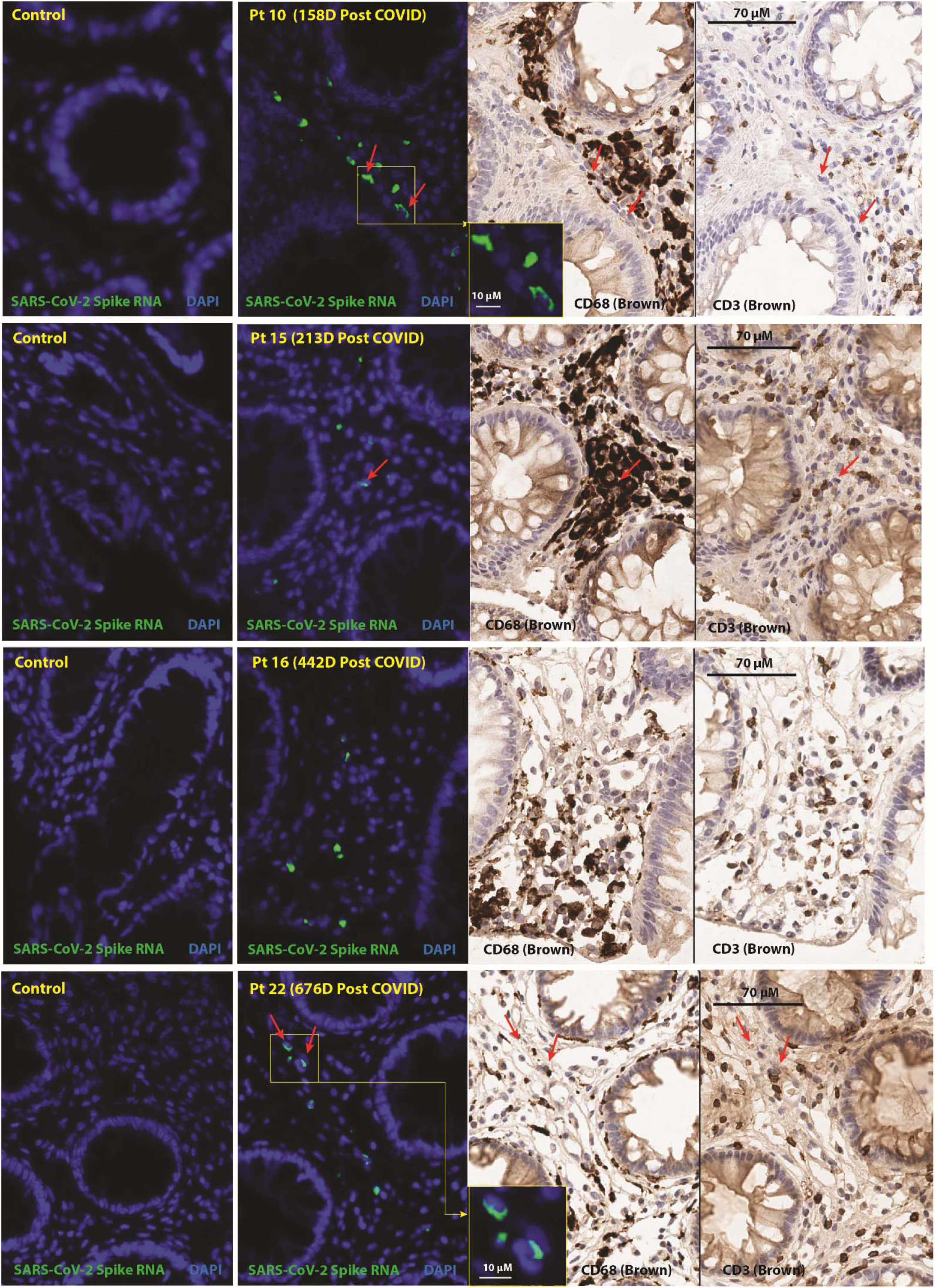
In situ hybridization (ISH) of SARS-CoV-2 spike RNA and immunohistochemical staining of recto-sigmoid tissue. Panels represent from left to right: SARS-CoV-2 spike RNA staining by ISH in pre-pandemic tissue, SARS-CoV-2 spike RNA staining by ISH in post-acute COVID participant sample, and CD68 and CD3 chromogenic immunostaining. Red arrows denote representative areas of RNA detection across images for each sample (not all RNA detection is marked). Spike RNA was detected in all five of the post-acute COVID participants that underwent biopsy from 158 to 676 days following initial COVID-19 symptom onset and signal was primarily observed in cells located within the lamina propria. Four participants with detectable RNA in three distinct gut regions are shown, a fifth participant had rare spike RNA detected in only one of three regions imaged. A minority of SARS-CoV-2 RNA signal was localized in CD68+ macrophages/immune cells and very rarely in CD3+ T cells. No viral RNA was detected in the pre-pandemic COVID tissue. Only linear adjustments to contrast and brightness were used during image analysis, with the same parameters being applied to all case and control tissues which underwent identical slide processing and imaging protocols.

### High-dimensional flow cytometric evaluation of peripheral blood and gut-derived mononuclear cell phenotypes

A multi-dimensional spectral flow cytometry panel that characterized CD4+ and CD8+ T cell, NK cell, and B cell phenotypes, including markers of activation, naive/memory phenotypes, regulatory function and immune checkpoint/exhaustion was performed on peripheral blood mononuclear cells (PBMC) from 16 participants who had sufficient PBMCs available from timepoints around the time of PET imaging and from gut tissue from 5 participants who underwent colorectal biopsies. Overall, we observed higher frequencies of effector memory CD8+ and CD4+ T cells in gut tissue versus peripheral blood, but similar frequencies of both CD8+ and CD4+ lymphocytes expressing activation markers CD38/HLA-DR and immune checkpoint (PD-1). In addition, there were no significant correlations between PET tissue uptake levels, Long COVID symptoms counts, timing of infection or vaccination or specific Long COVID symptom phenotypes with CD4+ and CD8+ T cell phenotypes (**Supplemental Figures 7 & 8**) or with early and late B cell memory phenotypes.

### Intestinal biopsies in a subset of post-acute COVID participants show evidence of persistent SARS-CoV-2 spike RNA in rectal tissue

Prior studies suggest that SARS-CoV-2 RNA or proteins may be detected in the gut or shed in stool for several months following acute COVID-19^34, 68^. Given the higher [^18^F]F-AraG uptake in proximal colon and rectal wall across many post-acute COVID participants compared with pre-pandemic controls, it is possible that viral persistence may be driving, at least in part, increased activated T cell migration to gastrointestinal tissues. We explored the potential for viral persistence by collecting rectosigmoid tissue by flexible sigmoidoscopy in a subset of 5 participants who had undergone PET imaging ranging from 158 to 676 days following initial infection (**Table 1**). Four of 5 participants had biopsies within 63 days of PET imaging; one participant (participant 14) was biopsied 182 days after imaging. All participants reported at least one Long COVID symptoms at the time of biopsy. None had received a SARS-CoV-2 vaccine dose in the prior month. None reported any history, symptoms, or testing suggestive of SARS-CoV-2 reinfection, and three of the five participants (participants 15, 16 and 18) who underwent rectosigmoid biopsy had no detectable SARS-CoV-2 nucleocapsid IgG detected within a short time frame of tissue collection (SC rations <1 at the time of PET imaging) suggesting no recent SARS-CoV-2 re-infection.

SARS-CoV-2 RNA was readily detected in multiple cells from all three tissue regions surveyed from all but one individual (participant 15) who had rare RNA+ cells detected in only one of three gut tissue regions 645 days following initial infection. However, another participant biopsied at a similar interval (participant 19) had higher spike RNA levels in gut tissue. Nearly all RNA was detected in cells in the lamina propria, without an epithelial signal (**Figure 6**). A small percentage of RNA+ cells expressed CD68, a macrophage monocyte lineage marker, but many RNA+ cells did not express CD68 and virtually none expressed CD3. [^18^F]F-AraG SUVmax values in proximal colon and rectal tissue in the post-acute COVID participants who underwent biopsy were at least 3 standard deviation above the mean SUVmax of pre-pandemic controls (1.86 vs 0.95 and 1.5 vs 0.9, respectively). PCR was also performed on RNA isolated from bulk rectal tissue lysates (separate biopsy from FFPE tissue used above) targeting the N1, N2, Envelope (E) and RNA-dependent RNA polymerase (RdRp) regions of SARS-CoV-2. No RNA was detected in any participant.

## DISCUSSION

In this first-in-human T cell activation PET imaging study of individuals in the post-acute phase of SARS-CoV-2 infection, we found that SARS-CoV-2 infection may result in persistent T cell activation in a variety of body tissues. In some individuals, this activity may persist for years following initial symptom onset and associate with systemic changes in immune activation as well as the presence of Long COVID symptoms. Finally, we found that SARS-CoV-2 persistence in gut tissue may contribute to these processes. Taken together, these observations suggest that even remote, clinically mild SARS-CoV-2 infection could have long-term consequences on tissue-based immune homeostasis. Our findings provide additional evidence to support the role of tissue-based immune activation and viral persistence as contributors to post-acute sequelae of SARS-CoV-2 infection, including Long COVID.

This study adds substantially to the existing literature on tissue-based immune responses in the post-acute phase of SARS-CoV-2 infection. Whereas traditional positron emission tomography (PET)-based imaging using radio-labeled glucose (FDG) as a marker of tissue inflammation has been applied to the study of acute and, to a lesser extent, post-acute COVID-19^46–52^, this method is non-specific because FDG is taken up by any metabolically active tissue. One pilot PET study using a CD8-specific minibody radiotracer provides some evidence that T cells traffic to tissues such as bone marrow following acute infection and remain up to 4 months thereafter^53^,https://paperpile.com/c/E2nfq2/yPPn but the relationship between T cell activation state, clinical symptoms, and viral persistence in the post-acute phase and over the years following COVID-19 was not addressed.

We found that [^18^F]F-AraG uptake was significantly higher in post-acute COVID participants compared to pre-pandemic controls in many anatomical regions, including brain stem, spinal cord, bone marrow, nasopharyngeal and hilar lymphoid tissue, cardiopulmonary vasculature, lung parenchyma, and gut wall. These observations were identified up to 2.5 years following initial COVID-19 symptom onset, in the absence of confirmed or suspected re-infection. Although [^18^F]F-AraG uptake in some tissues (spinal cord, colon/rectal wall) appeared to decline with time, the levels of uptake often remained elevated above those measured in pre-pandemic healthy control volunteers. These data significantly extend prior observations of a durable and dysfunctional cellular immune response to SARS-CoV-2^13, 21, 22, 69^ and suggest that SARS-CoV-2 infection could result in a new immunologic steady state in the years following COVID-19.

In this study, T cell activation in some tissues (spinal cord and gut wall) tended to be higher in participants reporting Long COVID symptoms of any type compared to both pre-pandemic controls and those post-COVID with complete recovery. Increased lung and hilar [^18^F]F-AraG uptake was identified in those with persistent pulmonary symptoms, suggesting a potential link between ongoing aberrant tissue immune responses and long-term clinical symptoms. It is now well-established that at least a subset of individuals with Long COVID exhibit prolonged systemic immune activation following SARS-CoV-2 infection^7, 10–21^. This study provides evidence for ongoing immune responses in tissues, a potentially important source of inflammation observed in peripheral blood. Furthermore, as Long COVID is increasingly being framed as having potential neurological underpinnings, it is possible that spinal cord and brainstem [^18^F]F-AraG uptake observed in our study may represent T cell trafficking to CNS tissues with residual viral components. This is consistent with a prior autopsy study, which demonstrated the presence of SARS-CoV-2 spike RNA and protein in the spinal cord and basal ganglia in two individuals that died during the post-acute phase following COVID-19 (65 and 230 days post infection)^38^.

We identified cellular SARS-CoV-2 RNA in rectosigmoid lamina propria tissue *in situ* in participants studied for up to two years following initial infection. This observation extends prior reports limited to 4-6 months post-COVID^34, 35, 38^. We employed several measures to guarantee virus-specific RNA *in situ* hybridization, including use of pre-pandemic control tissue on each slide to minimize technique differences or batch testing effects and repeated RNA staining in contiguous slices to verify consistency of location of the viral RNA signal. PCR from bulk tissue lysates was unsuccessful (and hence viral sequencing could not be performed), but SARS-CoV-2 RNAscope methods have been shown to be more sensitive in gut tissues in prior analyses^35^. As [^18^F]F-AraG uptake was observed in this anatomical region in nearly all participants, this finding suggests that virus persistence might contribute to the sustained levels of T cell activation observed in our participants. Because all of the participants who underwent gut biopsy met Long COVID criteria at the time of the procedure, it is difficult to draw concrete conclusions regarding the impact of gut viral persistence on Long COVID symptoms or [^18^F]F-AraG uptake. Further investigation, including assessment of fully recovered comparators, will be needed to determine whether persistence is clearly associated with Long COVID or occurs even in those who are believed to have fully recovered.

A key question for the field is in which cell type(s) SARS-CoV-2 might persist. We observed that some, but not a majority, of intracellular Spike RNA was associated with regions of CD68+ immune cell infiltrates (although not only within CD68+ cells), which likely represent tissue-resident macrophages within the lamina propria. ACE-2 expression in the lamina propria and on myeloid immune cells has been previously shown to be low, and we observed little-to-no RNA in the gut epithelium, where ACE-2 expression is higher^34, 70^. These data suggest that macrophages or other immune cells may be acquiring virus or viral contents either through phagocytosis of other infected cells or through viral-immune complexes as previously proposed^71^. Viral transcriptional activity may lead to innate immune sensing and downstream, tissue-based inflammation that could lead to infiltration of other immune cells (e.g., T cells), tissue damage and systemic inflammation even without replication or *de novo* infection.

Collectively, our data are consistent with a model in which persistence of SARS-CoV-2 results in chronic tissue-based inflammation, T cell activation and perhaps Long COVID. However, we acknowledge that this story of persistent immune activation and viral antigen in Long COVID is still developing. For example, we did not observe direct correlations between other Long COVID organ system-based phenotypes, such as gastrointestinal, neurologic, cardiac, and fatigue, and [^18^F]F-AraG uptake in anatomically related tissues. We also did not observe significant differences in overall T cell phenotypes in peripheral blood (memory subset, activation, exhaustion) between those with and without Long COVID or between participants with high or low [^18^F]F-AraG uptake in tissue ROIs in this cohort, in contrast to our previous observations in larger cohorts.^13, 21^ Our negative findings may be due to a smaller cohort or the well-known challenges in defining and measuring Long COVID^54^. Larger studies will be needed to further define the relationships between immune responses at the tissue level and specific Long COVID endotypes.

This study has several limitations, many inherent to PET imaging protocols. First, whereas the relatively small sample size limited power in certain tissue and immune correlative studies, we were powered to examine primary imaging endpoints (PET tracer uptake across post-acute COVID-19 cases and pre-pandemic controls). Second, because of the rapidly evolving pandemic, including intermittent variant waves, the recognition of Long COVID as a relevant clinical entity, and the rapid but inconsistent rollout of SARS-CoV-2 vaccines, we were required to adapt the protocol over time. This resulted in a shift from imaging participants closer to the time of initial SARS-CoV-2 symptom onset to prioritizing those with Long COVID symptoms months to years after COVID-19. As a result, those enrolled earlier in the pandemic tended to be imaged closer to the time of acute infection and were not selectively enrolled based upon the presence of Long COVID symptoms. Third, we relied mainly on pre-pandemic controls in healthy volunteers who were imaged in [^18^F]F-AraG protocols using similar PET acquisition strategies and timing from tracer injection to image acquisition. Pre-pandemic control volunteers were imaged using PET/MRI and, on average, received a higher dose of [^18^F]F-AraG, but SUV were used as comparison which take into account tracer injection dose, participant size and isotope decay rates and sensitivity of the PET-CT and PET/MRI scanners are similar. In addition, we would expect any confounding from this higher tracer dose to lead to higher uptake in the pre-pandemic controls compared to post-acute COVID participants. We note that it has become extraordinarily difficult to enroll contemporary, never-infected control participants in research studies now that most of the population has been either formally diagnosed or subclinically infected with SARS-CoV-2^72^. Finally, although mitigated by frequent assessments in the parent LIINC cohort, it remains possible that some participants may have had asymptomatic or undetected reinfections between the initial COVID-19 symptom onset and their rectosigmoidoscopy or [^18^F]F-AraG PET imaging.

In summary, our results provide provocative evidence of long-term immune system activation in several specific tissues following SARS-CoV-2 infection, including in those experiencing Long COVID symptoms. We identified that SARS-CoV-2 persistence is one potential driver of this ongoing activated immune state, and we show that SARS-CoV-2 RNA may persist in gut tissue for nearly 2 years after the initial infection. Overall, these observations challenge the paradigm that COVID-19 is a transient acute viral infection and provide evidence for T cell activation and viral persistence in tissues well beyond the initial illness.

## METHODS

### Participants

Study volunteers were participants in the University of California, San Francisco-based Long-term Impact of Infection with Novel Coronavirus (LIINC) study (NCT04362150^54^) and additionally opted into the imaging protocol (NCT04815096). Study procedures for the parent study have been described in detail elsewhere^54^. Briefly, adults with SARS-CoV-2 infection confirmed on nucleic acid-based or antigen-based testing were enrolled 14 days or longer following initial COVID-19 symptom onset and assessed prospectively approximately every 4 months following initial symptom onset. Participants were excluded from the imaging protocol if they had a history of excessive radiation, underwent a prior research study involving radiation within 1 year of enrollment, were pregnant or breastfeeding, had screening absolute neutrophil count <1000 cells/mm3, platelet count <75,000 cells/mm3, hemoglobin <8 g/dL, estimated creatinine clearance <60 mL/min, aspartate aminotransferase >3x ULN units/L, alanine aminotransferase >3x ULN units/L, had recent use of medication including guanosine or cysteine analogs, had known SARS-CoV-2 nasopharyngeal shedding within 5 days of scan, had a SARS-CoV-2 vaccine within 4 weeks of scan, or had a prior history of immunoproliferative or autoimmune disease.

### Questionnaire-based measurements

At the first visit, participants completed an interviewer-administered questionnaire that assessed demographics, medical history, and SARS-CoV-2 infection, vaccination, and treatment history. At all visits, participants completed additional interview-administered questionnaires that queried the presence and severity of any symptoms that were new or worsened since the initial SARS-CoV-2 diagnosis, quality of life, interim medical diagnoses, and interim treatments and vaccinations. Symptoms that predated SARS-CoV-2 infection and were unchanged were not considered to represent Long COVID. Concurrent with PET imaging, detailed histories were obtained including symptoms, prior or subsequent SARS-CoV-2 PCR or antigen testing, and clinical symptoms potentially related to SARS-CoV-2 re-infection.

### [^18^F]F-AraG imaging

Following study entry and eligibility screening, participants were administered intravenously [^18^F]F-AraG (166.5-185 MBq) and PET/CT whole-body imaging was carried out for 20 min at approximately 50 min post injection. [^18^F]F-AraG was prepared as documented elsewhere (15). Images were taken from the top of head to mid thighs. Participants were observed at imaging visit or reported by participants after [^18^F]F-AraG imaging per phone calls by study coordinators 2, 7, and 30 days after imaging. Routine urinalysis was performed 7-14 days after imaging to ensure proper excretion of [^18^F]F-AraG.

### PET and CT image analysis

Standardized uptake values (SUV) in various tissue regions of interest (ROI) from PET/CT data were determined using the OsiriX DICOM viewer software package (Pixmeo; Bernex Switzerland). ROI determination was performed in complex structures such as brain sections, heart wall, spleen, and gut wall using two-dimensional isometric ROIs. For simpler structures such as the spinal cord at various levels, bone marrow, and whole lymph nodes, three-dimensional spherical ROIs were used. For axillary and inguinal lymph node ROI selection, the most prominent nodes on both the right and left side were included and SUV values averaged. ROI selection was performed independently by two individuals blinded to the study group following ROI selection upon a subcohort and comparison to ensure consistency across reviewers. Two-and three-dimensional PET or PET/CT images were generated in OsiriX keeping window levels consistent between participants. SUVs from participants reporting PASC symptoms were compared with six pre-pandemic PET/MRI controls and, in two post-pandemic (contemporary) PET/CT controls. ROI selection on bowel tissue was challenging as we observed intermittent intraluminal collection of tracer, which was highly anatomically variable across all participants. As intraluminal signal does not represent specific tissue tracer uptake, ROI selection was performed in gut wall tissue only in areas without clear contiguous intraluminal signal. As a result, ROI selection was not possible in 3 and 5 post-acute COVID participants for proximal colon and rectal wall ROIs, respectively, and 3 pre-pandemic control participants for proximal colon. All PET images and CT images of the chest were further reviewed independently by two dual board-certified radiologists and nuclear medicine physicians (R.R.F. and Y.W.). Qualitative abnormalities were reviewed and tabulated.

### Circulating Markers of Inflammation

A Protein Extension Assay (PEA) using the Olink EXPLORE Inflammation panel from plasma samples was performed in post-acute COVID imaging participants to characterize 365 unique plasma proteins associated with inflammation and immune signaling. Briefly, PEA involves dual-recognition of two matched antibodies labeled with unique DNA oligonucleotides that simultaneously bind to specific target proteins. The simultaneous antibody binding leads to hybridization of unique DNA oligonucleotides that serve as templates for polymerase-dependent extension (DNA barcoding) followed by PCR amplification and NovaSeq (Illumina) DNA sequencing. Clustered heatmaps were generated by the UCSF Gladstone Bioinformatics Core using the R package HOPACH to find the best cluster number. Gene product expression values were log-transformed and centered using the average expression value for each protein. Gene products were then clustered by running the Kmeans algorithm using the best cluster number K found, and the results were plotted using the pheatmap package as modules. Standard deviation (SD) cutoff levels between those with higher and lower [^18^F]F-AraG uptake for each tissue were decided based on variance of SUVmax values of uninfected controls, with higher variance allowing for one or more SDs above the mean to qualify as “higher” uptake.

### SARS-CoV-2 qPCR on Rectal Biopsies, PCR and In situ hybridization of SARS-CoV-2 Spike RNA

Rectal wall tissue samples were obtained via flexible sigmoidoscopy with tissue being fixed in fresh paraformaldehyde (PFA) followed by Paraffin embedding approximately 48 hours after fixation or were cryopreserved at −180°C in Fetal Bovine Serum (FBS) and 20% Dimethyl sulfoxide (DMSO) as described^73^. A minimum of three formalin-fixed paraffin embedded (FFPE) rectosigmoid tissue biopsies from each participant were used for RNAscope experiments, along with comparative uninfected tissue, were mounted on the same slide to control for batch effects from processing, staining, microscopy and image analysis. Experiments were performed at least twice on contiguous sections to verify signal over non-specific staining or autofluorescence. Contiguous sections underwent hematoxylin and eosin staining and immunohistochemical visualization of CD3 and CD68 expression in order to localize viral RNA signals with anatomical tissue regions and immune cell types of interest. Quantitative PCR assays were performed using Integrated DNA Technology’s (IDT) SARS-CoV-2 RUO qPCR Primer & Probe Kits for N1, N2, E, and RDRP detection (catalog no. 10006713, 10006804, 10006805, and 10006806). Positive controls consisted of fragments of human RPP30 and SARS-CoV-2 isolate Wuhan-Hu-1 (GenBank: NC_045512.2) provided in each IDT kit. Quantitative PCR was performed with TaqPath™ 1-Step RT-qPCR Master Mix with the following conditions: 95 °C 2 min, 95 °C 3 sec, 55 °C 30 sec using the StepOnePlus™ Real-Time PCR System. SARS-Cov-2 RNA was considered detectable for cycle threshold (ct) values <40. Positive and non-template controls were run for all samples tested.

The manual RNAscope 2.5 HD assay (Advanced Cell Diagnostics; catalog no. 322310) was used to identify SARS-CoV-2 Spike RNA *in situ*. Paraffin-embedded tissue blocks were sectioned at 5 μm, mounted onto SuperFrost Plus slides, and stored at 4°C prior to staining. Slides were baked in a dry-air oven at 60°C for 1 h, then deparaffinized in 100% Xylene (5 min) twice and washed in 95% ethanol (3 min) twice, 80% ethanol (3 min) once, and dH2O (1 min) twice, all at room temperature. To prevent drying, 3-4 drops of TBS were placed on each tissue section. A hydrophobic barrier was then drawn around each tissue section and allowed to dry for 10 min. To block endogenous peroxidase slides were then pretreated with hydrogen peroxide for 10 mins at room temperature followed by washing with dH2O. Next, heat-induced epitope retrieval was performed with Target Retrieval Reagent (ACDBio) and incubated at 100°C for 15 minutes followed by rinsing with dH2O. Protease digestion was accomplished by treatment with Protease Plus solution (ACDBio) for 30 minutes at 40°C followed by dH2O wash. Hybridization was performed with RNAscope™ probe-V-nCoV2019-S (catalog no. 848561-C3, Advanced Cell Diagnostics) at 40°C for 2 h at a 1:50 dilution. Following hybridization, 3 amplification steps were carried out as indicated in the original protocol. Slides were then incubated with HRP-C3 at room temperature for 15 min, TSA Vivid Fluorophore 520 for 40°C for 30 min, and HRP blocker at 40°C for 15 min. Finally, slides were counterstained with DAPI, washed in PBS, and cover-slipped using Prolong Diamond Mounting Media.

Images were captured using the Zeiss AxioObserver Z1 (RNAscope) or the Leica Aperio VERSA (chromogenic staining on contiguous tissue sections). FFPE rectal tissue from a pre-pandemic control participant was mounted on each participant slide to control for any staining or imaging technique differences. Only linear brightness and contrast adjustments were made to image files and all adjustments were applied identically for each image.

### Spectral Flow Cytometry

PBMC or cells obtained from gut tissue following a previously published collagenase disaggregation protocol^74^ were Stained with the Cytek 25-plex kit (Cat# SKU R7-40002) with the BioLegend antibodies (Cat#900004160), for Live-dead staining Zombie UV (Cat#423108) was used. Cells were stained with titrated antibodies for 30 min at 25 degrees Celsius in a total volume of 130ul, including the addition of Brilliant stain buffer plus (cat#566385). Cells were washed twice with FACS buffer (PBS+10%FBS+1mM EDTA) before fixation with 1% PFA (in PBS). Cells were acquired on the 5L Cytek Aurora the day after. We used Spectrflow beads to verify laser alignment and power consistency. The forward and side scatter profiles were established using human PBMCs, similar to the test samples. Single stained cellular reference controls were used to unmix the data according to the 25-plex Cytek acquisition protocol, with unstained cells as an autofluorescence control. Unmixing errors were corrected by spillover correction using OMIP-069 supplementary information as a guide. The Cytek analysis template was used to replicate the gating strategy in FlowJo 10 as shown in **Supplemental Figures 9 & 10**. Data was exported, and further analysis performed on GraphPad Prism.

### SARS-CoV-2 Nucleocapsid Antibody Testing

Nucleocapsid IgG antibodies from post-acute COVID participants from plasma tested at the time of PET imaging were measured using the Abbott Architect i2000 two step Chemiluminescent microparticle immunoassay (CMIA). Signal to cutoff (S/C) rations were determined and samples were considered positive if the S/C ratio was above the assay-defined threshold (S/C >1 were considered positive).

### Statistical analysis

We used two-tailed, non-parametric Kruskal-Wallis tests using a Benjamini-Hochberg adjustment for false discovery rates from multiple comparisons within specific tissue regions (e.g. lymphoid tissues, glandular tissue, vascular, spinal cord, etc.) to compare ROI SUV data and flow cytometric data across participants (adjusted P values being analogous to the Q value obtained from FPR adjustments). Nonparametric tests were used given the assumption that SUV data may not be normally distributed across comparator groups. Two-sided Spearman rank tests were performed to determine correlations between continuous variables.

### Human subjects

Participants provided written informed consent for both the parent study and the imaging protocol. The study was approved by the UCSF Institutional Review Board and the UCSF Radiation Safety Committee. ClinicalTirals.gov numbers: NCT04362150 and NCT04815096.

## FOOTNOTES

## Data Availability

All data produced in the present study are available upon reasonable request to the authors

## Acknowledgements

We are grateful to the study participants and their medical providers. We acknowledge current and former LIINC clinical study team members Tamara Abualhsan, Andrea Alvarez, Grace Anderson, Khamal Anglin, Urania Argueta, Mireya Arreguin, Alexus Clark, Nicole DelCastillo, Emily Fehrman, Halle Grebe, Heather Hartig, Yanel Hernandez, Beatrice Huang, Marian Kerbleski, Raushun Kirtikar, Suzanna Kouzi, Megan Lew, James Lombardo, Monica Lopez, Michael Luna, Lynn Ngo, Enrique Martinez Ortiz, Justin Romero, Ruth Diaz Sanchez, Matthew So, Celina Chang Song, Alex Tang, Cassandra Thanh, Fatima Ticas, Leonel Torres, Brandon Tran, Daisy Valdivieso, Deepshika Varma, Meghann Williams, and Andhy Zamora; and LIINC laboratory team members Joanna Donatelli, Jill Hakim, Nikita Iyer, Owen Janson, and Keirstinne Turcios. We thank Jessica Chen, Aidan Donovan, Carrie Forman, Rania Ibrahim, and Badri Viswanathan for assistance with data entry and review. We thank the UCSF AIDS Specimen Bank for processing specimens and maintaining the LIINC biospecimen repository. We are grateful to Elnaz Eilkhani for regulatory support. We are also grateful for the contributions of additional LIINC leadership team members: Bryan Greenhouse, Isabelle Rodriguez-Barraquer, and Rachel Rutishauser.

## Author Contributions

Study design: MJP, DR, RF, HV, JL, AG, TJH

Acquired funding: TJH, MJP, HV, SGD

Parent Cohort Design and Oversight: MJP, RH, MSD, PYH, JDK, JNM, SDG, TJH

Participant Recruitment and Clinical Data Collection: MJP, DR, KA, MA, WK, SEM, RH, MD, AR, MB, VT, TJH

Data management and curation: SL, SAG, TD

Gut Tissue Collection: MD, AR, MS

Assay Design: DR, BL, BL, GS, ZGL, NK, TJH

Laboratory and Data Analysis: BHL, TMD, KA, SEM, GS, ZGL, NK, TJH

Image Analysis: DR, RF, YW, JL, KA, US, YS, TJH

Manuscript Writing: MJP, DR, RF, SGD, HV, TJH

Manuscript editing/reviewing: All authors

## SUPPLEMENTAL INFORMATION

**Supplemental Table 1.** Clinical Findings on Chest CT Scans.

| Pt # | Days from Infection to PET Scan<sup>a</sup> | Sex | Age Bracket (Years) | Hosp | LC Symptom Count<sup>b</sup> | Abnormal Findings on Chest CT as Determined by Two Cross-Sectional Radiologists (R1, R2) |
| --- | --- | --- | --- | --- | --- | --- |
| 1 | 27 | F | 25-29 | N | 2 | None |
| 2 | 29 | M | 35-39 | N | 1 | R Lower Lobe Bulla (R1); Minimal basilar subpleural bands/reticulation/ground glass (R2) |
| 3 | 42 | M | 30-34 | N | 4 | None |
| 4 | 44 | M | 60-64 | N | 0 | Minimal subpleural reticulation and banding in bases, L>R (R2) |
| 5 | 48 | M | 25-29 | N | 5 | None |
| 6 | 50 | F | 30-34 | N | 4 | Few tiny subpleural micronodules, likely intrapulmonary lymph nodes (R2) |
| 7 | 64 | M | 40-44 | N | 14 | None |
| 8 | 65 | F | 60-64 | N | 0 | Mild apical scarring |
| 9 | 83 | F | 55-59 | N | 0 | None |
| 10 | 95 | M | 45-49 | N | 0 | Calcified R lower lobe granuloma, L lower lobe subpleural granuloma (R1) |
| 11 | 114 | F | 30-34 | N | 8 | Subpleural L lower lobe nodule, intrapulmonary lymph node (R1) |
| 12 | 193 | F | 25-29 | N | 13 | None |
| 13 | 205 | F | 45-49 | N | 6 | Minimal apical, lingular and R middle lobe consolidation or scarring (R1) |
| 14 | 231 | M | 35-39 | N | 4 | None |
| 15 | 246 | M | 65-69 | N | 6 | None |
| 16 | 260 | M | 50-54 | Y<sup>c</sup> | 8 | R lower lobe nodule <1 centimeter (R1 & R2) |
| 17 | 406 | F | 30-34 | N | 7 | L major fissure micronodule, intrapulmonary lymph node (R1) |
| 18 | 617 | F | 30-34 | N | 14 | None |
| 19 | 625 | M | 45-49 | N | 11 | None |
| 20 | 641 | M | 25-29 | N | 15 | None |
| 21 | 654 | F | 30-34 | N | 6 | None |
| 22 | 663 | M | 50-54 | N | 0 | Mild apical scarring |
| 23 | 890 | M | 50-54 | N | 6 | Basilar predominant subpleural reticulation and scarring |
| 24 | 910 | F | 55-59 | Y<sup>d</sup> | 0 | None |

R = right; L = left; M = male; F = female
<sup>a</sup> Days from last COVID-19 vaccine dose to PET imaging
<sup>b</sup> Number of participant reported symptoms at the time of PET imaging (out of 32 total)
<sup>c</sup> Participant did not require intensive care but received supplemental oxygen during hospitalization
<sup>d</sup> Participant did not require intensive care or supplemental oxygen during hospitalization

**Supplemental Figure 1.**
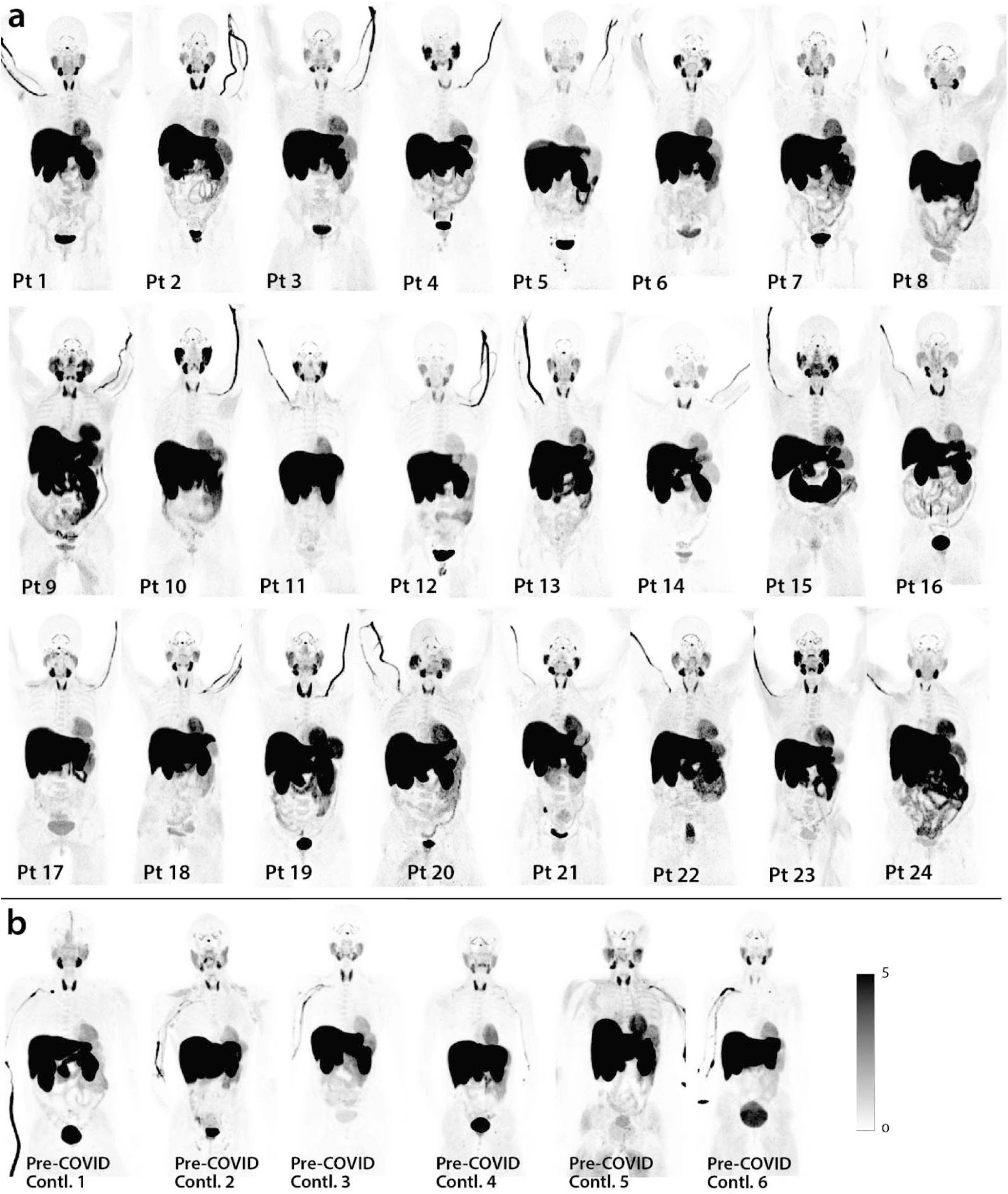
Maximum intensity projections (MPI; coronal views of 3-dimesional reconstructions) are shown for all cases and pre-pandemic and contemporary control participants.

**Supplemental Figure 2.**
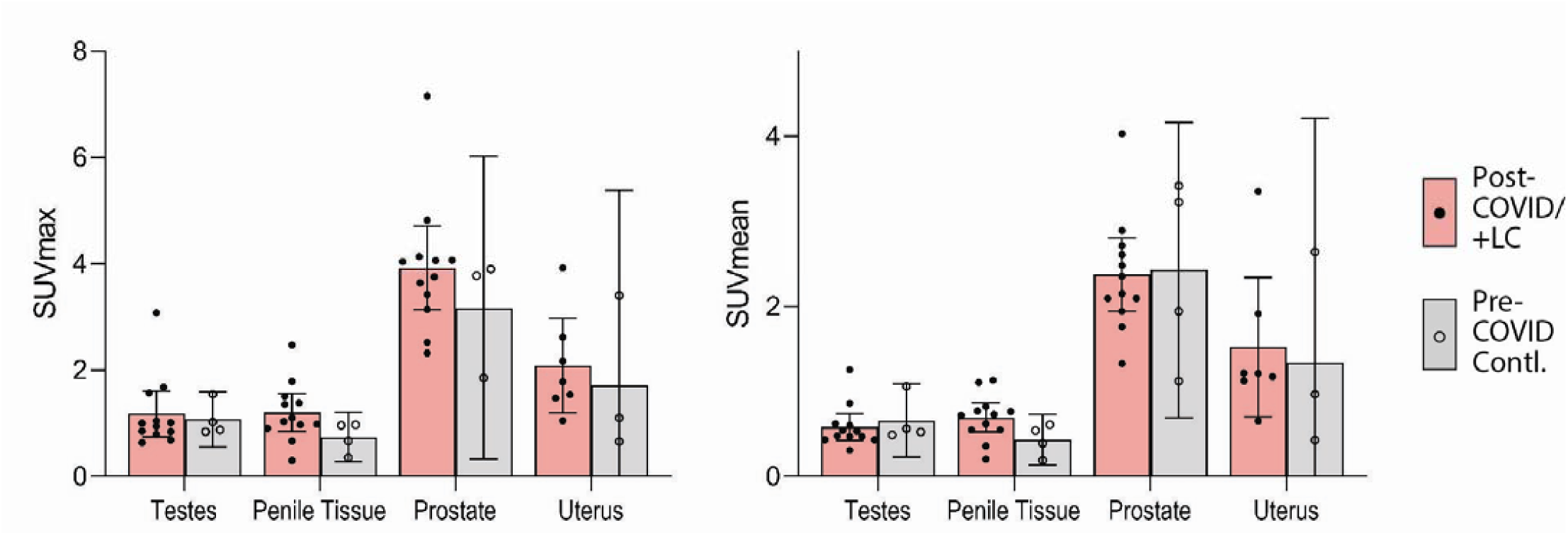
[^18^F]F-AraG SUVmax and SUVmean for reproductive tissues for cases and pre-pandemic controls are shown. Bars represent mean SUVmax and error bars represent 95% confidence interval. No statistical differences were observed between cases and controls for any reproductive tissue ROI. All data points are shown.

**Supplemental Figure 3.**
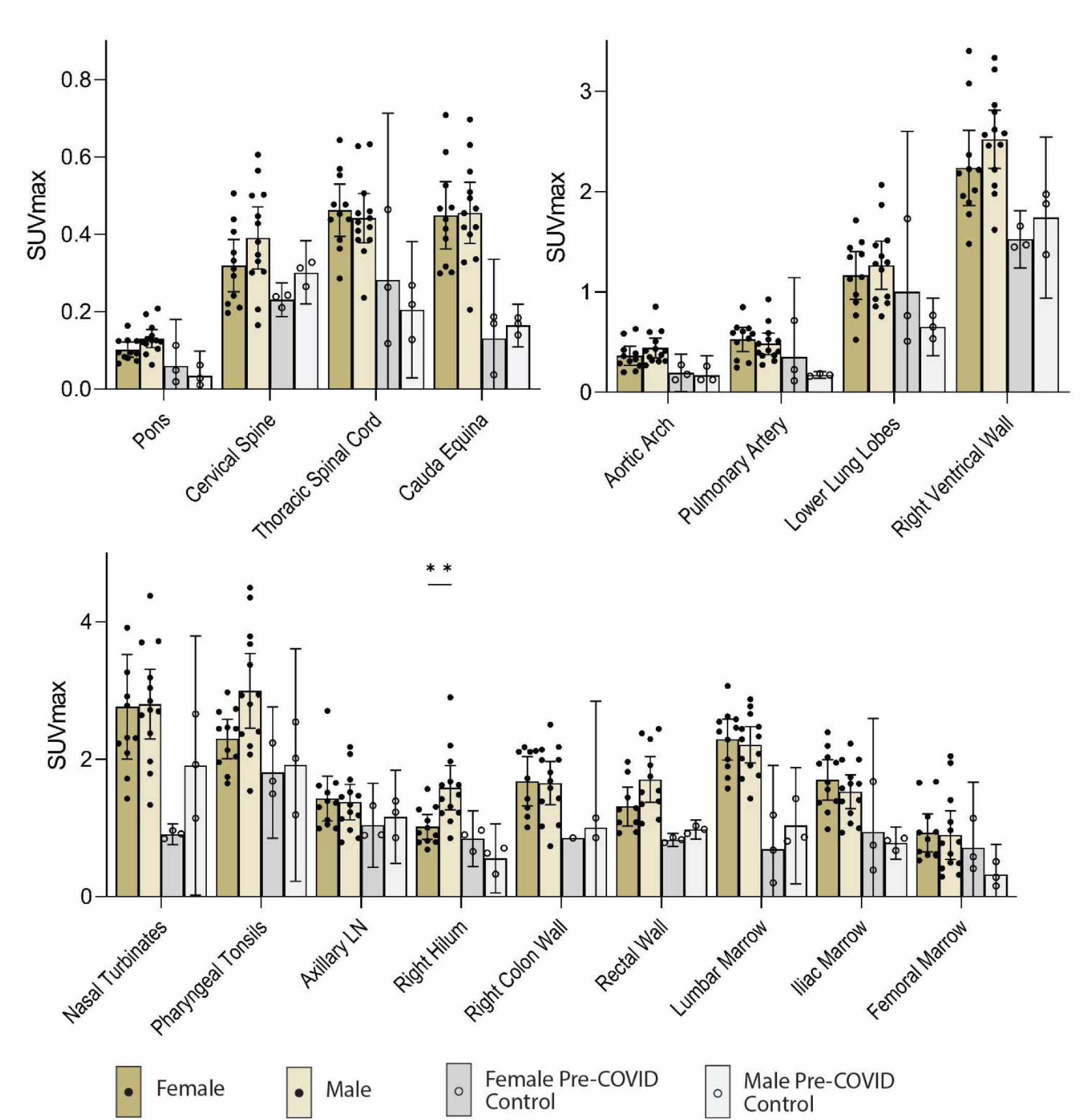
Comparisons of [^18^F]F-AraG maximum standardized uptake values in post-acute COVID cases and pre-pandemic control participants grouped by sex assigned at birth are shown. Bars represent mean SUVmax and error bars represent 95% confidence interval. Adjusted P values <0.05 and <0.001 represented by * and **, respectively from two-sided non-parametric Kruskal–Wallis tests using a Benjamini-Hochberg adjustment for false discovery rates across multiple comparisons (q value = adjusted P). Given lack of power to compare cases versus controls grouped on sex, statistical analyses were only performed between female and male post-acute COVID participants. All data points are shown.

**Supplemental Figure 4.**
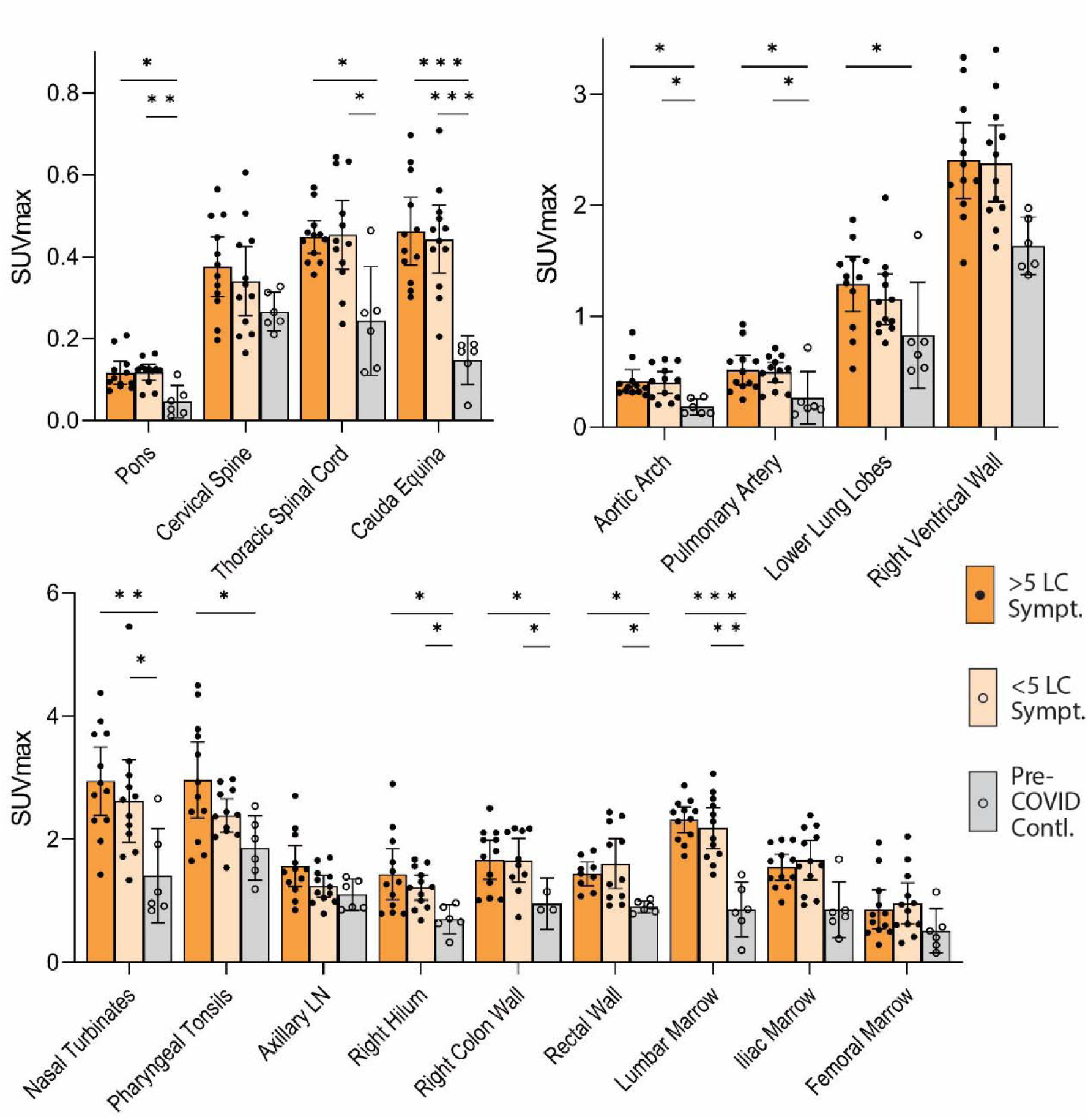
Comparisons of [^18^F]F-AraG maximum standardized uptake values in post-acute COVID cases and pre-pandemic control participants grouped participants with >5 or ≤5 Long COVID symptoms reported at the time of imaging and control volunteers are shown. Bars represent mean SUVmax and error bars represent 95% confidence interval. Adjusted P values <0.05, <0.01 and <0.001 represented by *, **, and *** respectively from two-sided non-parametric Kruskal–Wallis tests using a Benjamini-Hochberg adjustment for false discovery rates across multiple comparisons (q value = adjusted P). All data points are shown.

**Supplemental Figure 5.**
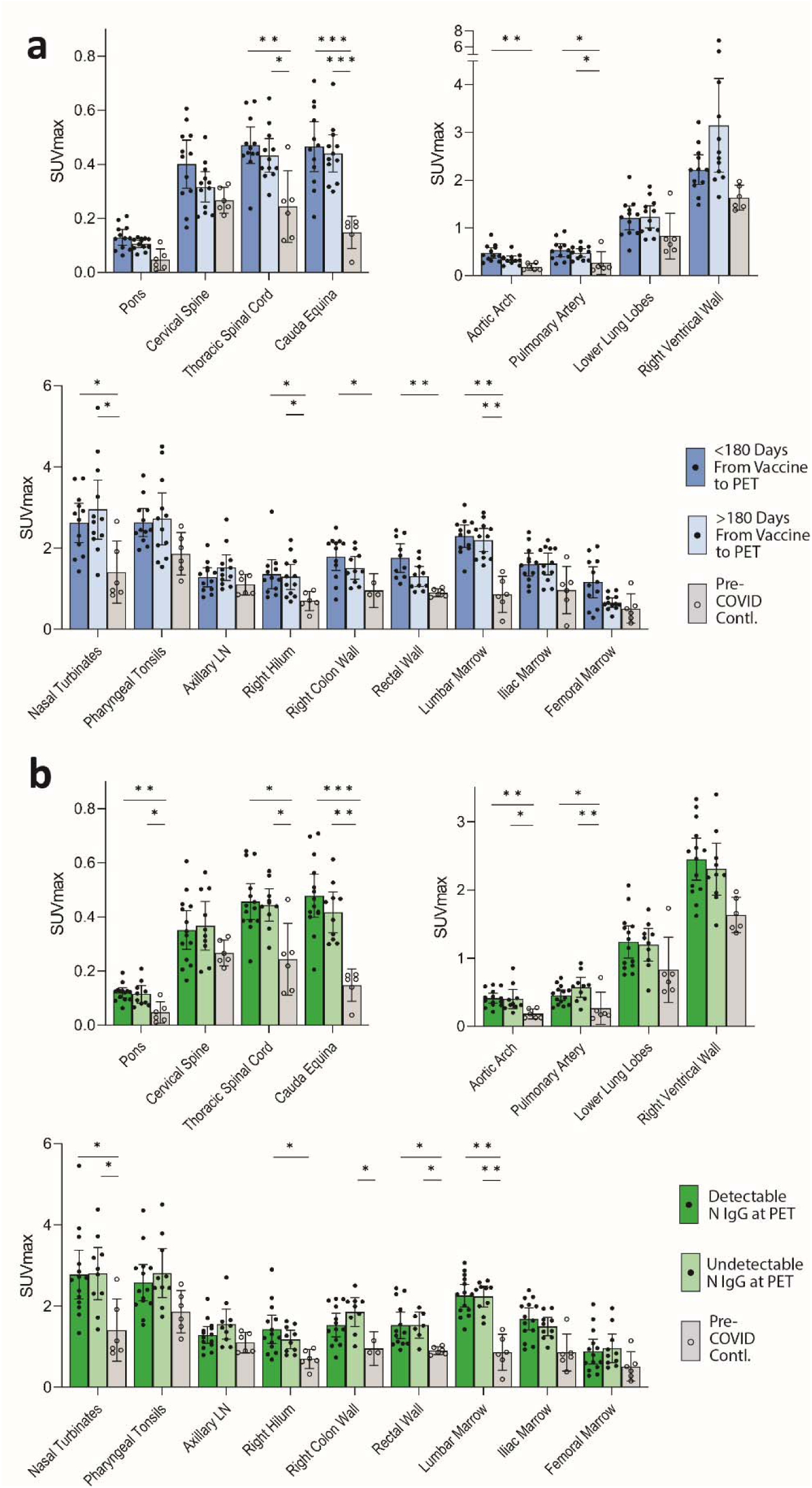
Comparisons of [^18^F]F-AraG maximum standardized uptake values in post-acute COVID cases and pre-pandemic control participants grouped by time from most recent dose of SARS-CoV-2 vaccine and PET imaging and by presence of detectable nucleocapsid (N) IgG detection at the time of imaging. SUVmax values in tissue ROIs in post-acute COVID participants imaged <180 days or >180 days from the last dose of COVID-19 vaccine and control volunteers are shown in (**a**). SUVmax values in tissue ROIs in post-acute COVID participants with detectable or undetectable SARS-CoV-2 N IgG measures at the time of imaging and control volunteers are shown in (**b**). Bars represent mean SUVmax and error bars represent 95% confidence interval. Adjusted P values <0.05, <0.01 and <0.001 represented by *, **, and *** respectively from two-sided non-parametric Kruskal–Wallis tests using a Benjamini-Hochberg adjustment for false discovery rates across multiple comparisons (q value = adjusted P). All data points are shown.

**Supplemental Figure 6.**
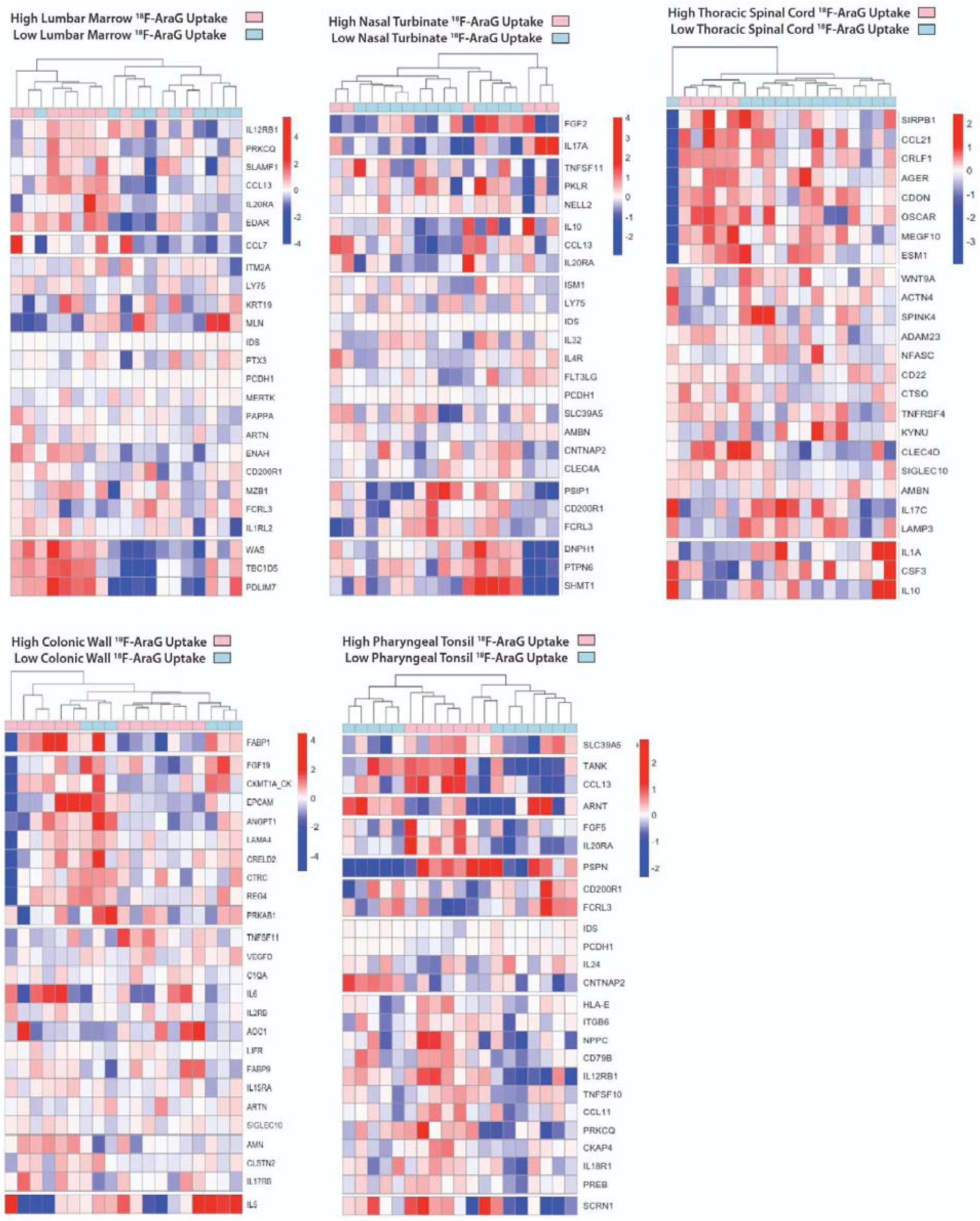
Differential plasma protein expression in post-acute COVID participants grouped by high or low [^18^F]F-AraG uptake in representative tissues. Clustered heat maps of the top 25 differentially expressed plasma proteins from Olink Proximity Extension Assay EXPLORE 384 panel with markers grouped into k-clusters based on similarity are shown for participants with high or lower PET signal in various tissue ROIs.

**Supplemental Figure 7.**
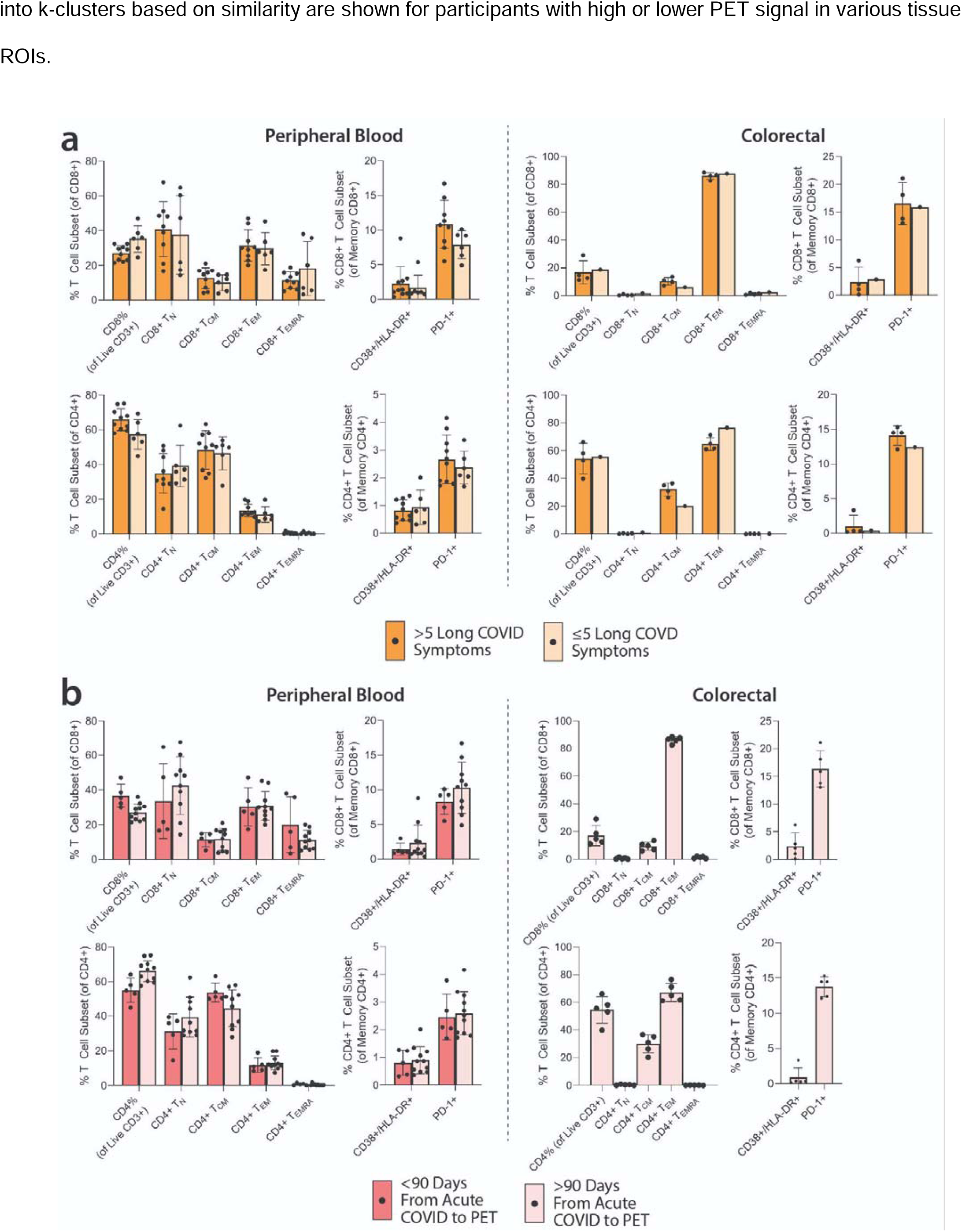
Spectral flow cytometry results for T lymphocyte phenotypes. The frequency of CD8+ and CD4+ T cell subsets (TN = naïve, TCM = central memory, TEM = effector memory, TEMRA = effector memory RA+/terminally differentiated) and frequency of lymphocytes co-expressing activation markers CD38/HLA-DR and immune checkpoint PD-1 from peripheral blood and gut for those with >5 or ≤5 Long COVID symptoms (**a**) and those imaged >90 or <90 days after onset of acute COVID-19 (**b**) are shown. No significant differences between post-acute COVID groups were identified. Bars represent percent of T cells expressing markers of interest and error bars represent 95% confidence intervals.

**Supplemental Figure 8.**
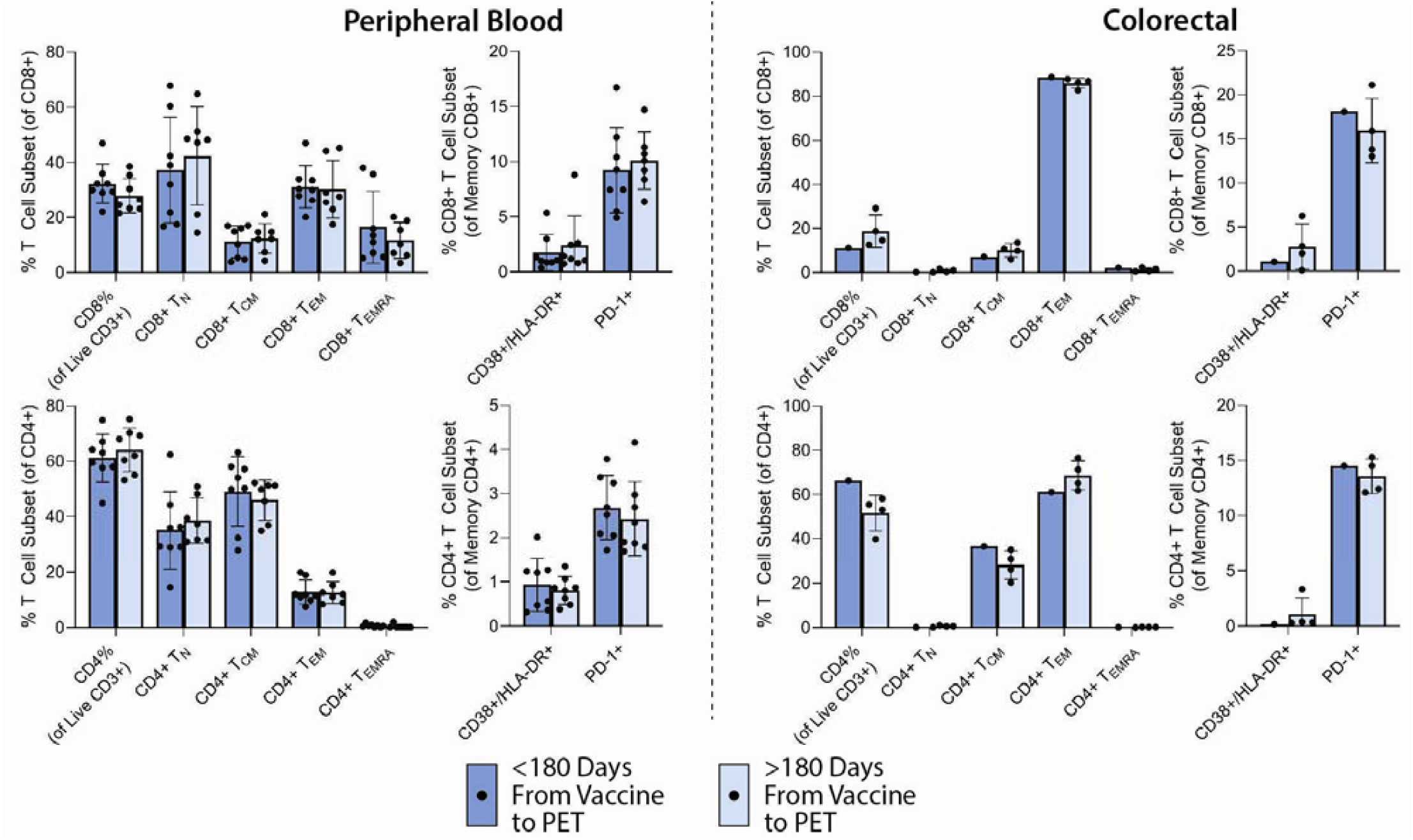
Spectral flow cytometry results for T lymphocyte phenotypes grouped by time of PET imaging from last COVID-19 vaccine dose. The frequency of CD8+ and CD4+ T cell subsets (TN = naïve, TCM = central memory, TEM = effector memory, TEMRA = effector memory RA+/terminally differentiated) and frequency of lymphocytes co-expressing activation markers CD38/HLA-DR and immune checkpoint PD-1 from peripheral blood and gut for those who underwent PET imaging <180 or >180 days following the last dose of COVID-19 vaccine are shown. No significant differences between post-acute COVID groups were identified. Bars represent percent of T cells expressing markers of interest and error bars represent 95% confidence intervals.

**Supplemental Figure 9.**
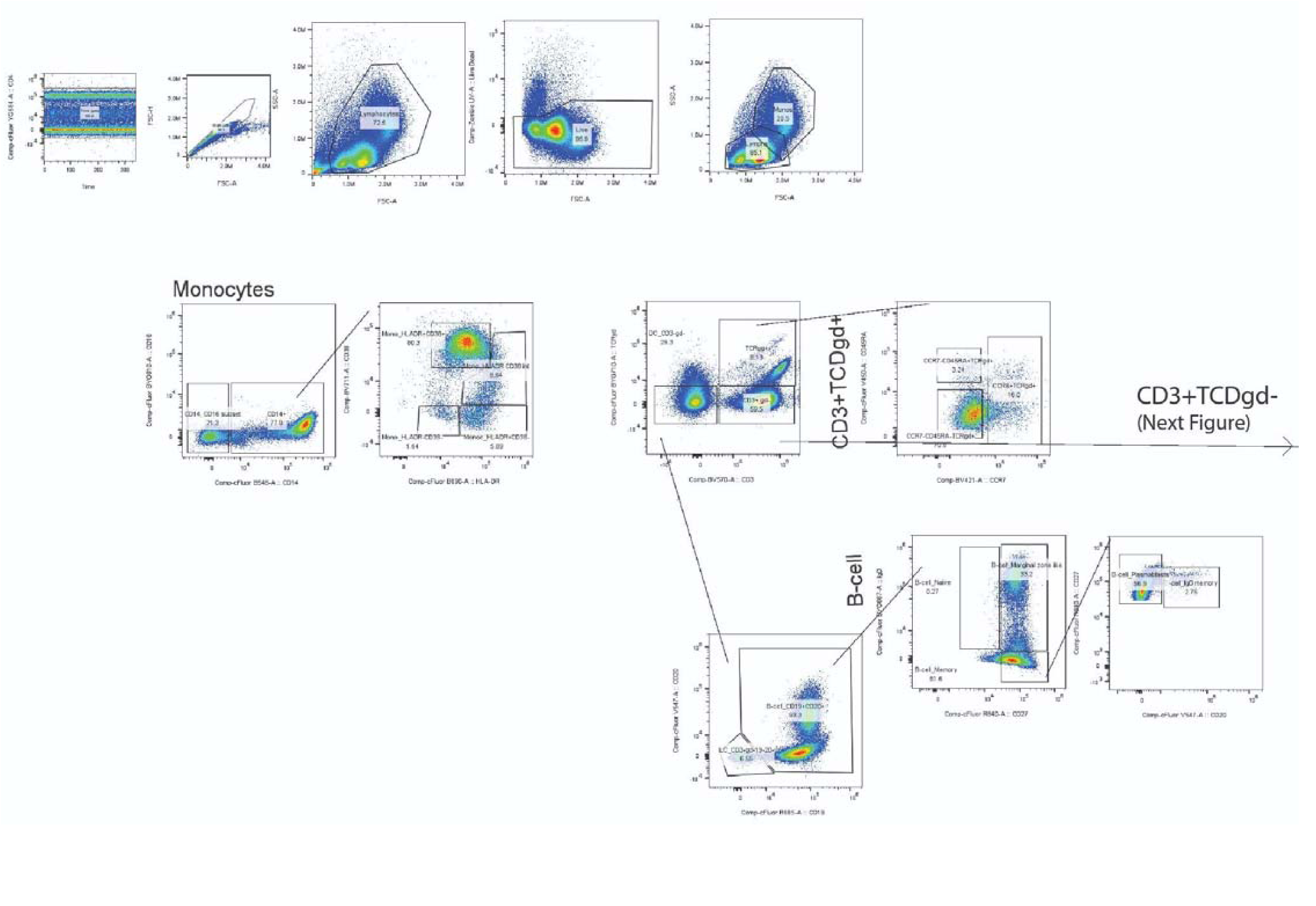
Lymphocyte and other mononuclear cell gating strategy (part A).

**Supplemental Figure 10.**
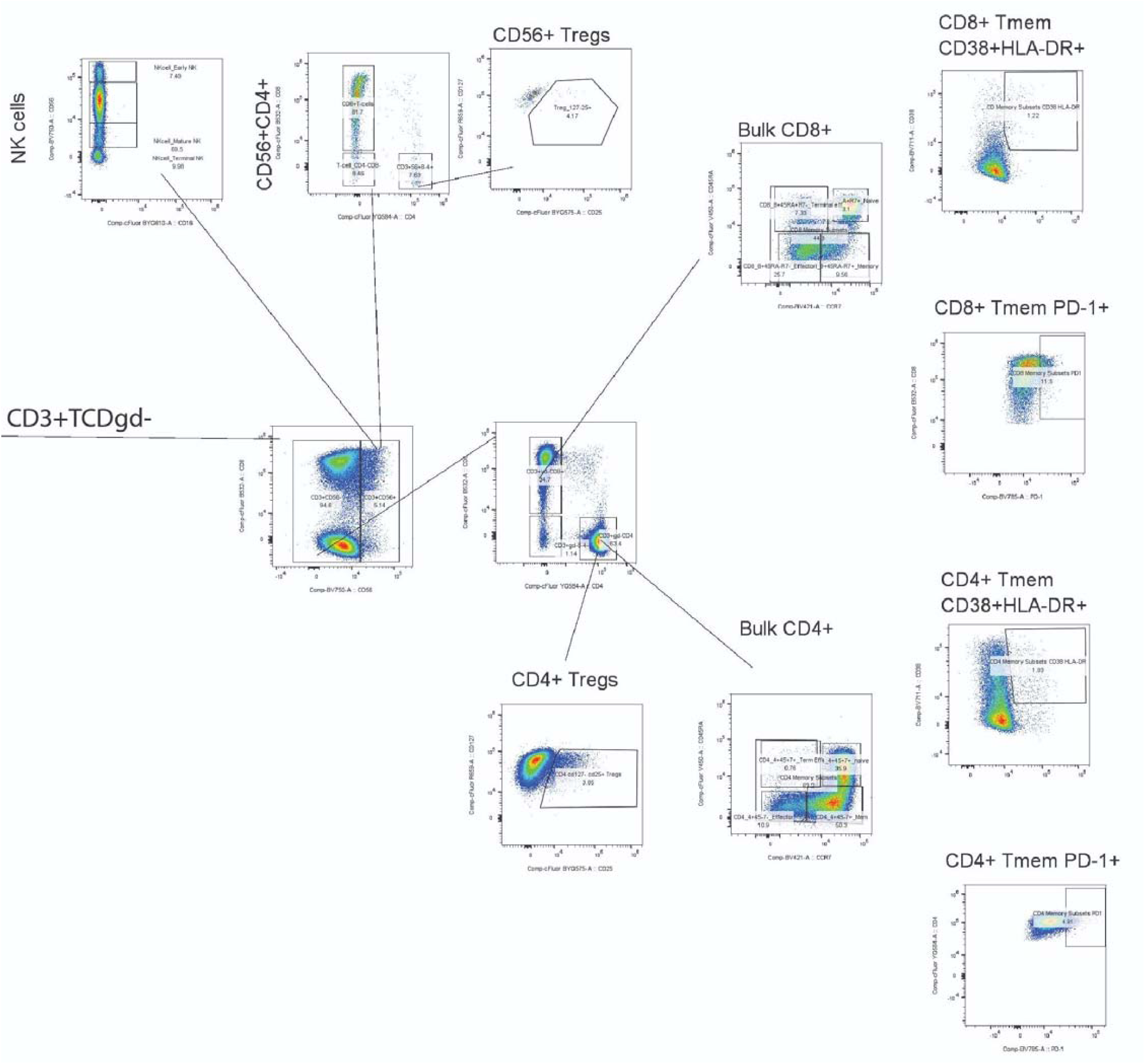
Lymphocyte and other mononuclear cell gating strategy (part B).

